# Cell-free DNA Whole Genome Sequencing for Non-Invasive Minimal Residual Disease Detection in Multiple Myeloma

**DOI:** 10.1101/2025.10.24.25338566

**Authors:** Dor D. Abelman, Jenna Eagles, Aimee Wong, Saumil Shah, Jeff P. Bruce, Stephanie Pedersen, David S. Scott, Cecilia Bonolo de Campos, Signy Chow, Ellen N. Wei, Safa Abdulsalam, Darrell White, Irwindeep Sandhu, Kevin Song, Esteban Braggio, Shaji Kumar, Alli Murugesan, Anthony Reiman, A. Keith Stewart, Suzanne Trudel, Trevor J. Pugh

**Affiliations:** Department of Medical Biophysics, University of Toronto; Princess Margaret Cancer Centre, UHN; School of Integrated Health, University of New Brunswick, Saint John; Faculty of Medicine, Dalhousie University; Odette Cancer Centre and Sunnybrook Health Sciences Centre; Division of Medical Oncology and Hematology, Temerty Faculty of Medicine, University of Toronto; Queen Elizabeth II Health Sciences Centre; Cross Cancer Institute; Vancouver General Hospital; Department of Cancer Biology, Mayo Clinic Arizona; Mayo Clinic Rochester; Faculty of Science, Applied Science and Engineering, University of New Brunswick, Saint John; Department of Oncology Saint John Regional Hospital; Ontario Institute for Cancer Research

## Abstract

Minimal residual disease (MRD) monitoring in multiple myeloma (MM) relies on invasive bone marrow (BM) biopsies, which often yield insufficient tumor material. We performed whole genome sequencing of cell-free DNA from 163 plasma samples from 51 patients to develop a non-invasive MRD classifier. BM WGS identified a median of 2,502 clonal mutations, enabling cfDNA tracking at levels comparable to BM-based MRD testing. The cfDNA classifier achieved a mean AUC of 0.86 against multiparameter flow cytometry and targeted immunoglobulin sequencing (clonoSEQ), and MRD negativity after one year of maintenance was strongly associated with two-year relapse-free survival probability (Hazard Ratio = 24), with cfDNA changes preceding clinical progression by a median of 12.6 months. To establish a BM-agnostic mode, a plasma-only classifier trained on baseline cfDNA established a mean AUC of 0.79 compared to BM clinical testing and stratified relapse risk (HR = 4.18), enabling MRD detection in patients with suboptimal BM samples. Longitudinal cfDNA profiles detected subclonal evolution in half of profiled patients. Cell-free DNA whole genome sequencing provides a sensitive, scalable, and clinically informative platform for non-invasive MRD monitoring in MM.

**Statement of Translational Significance:** Currently, myeloma MRD assays require invasive, painful bone marrow sampling and fail to account for spatial heterogeneity. High-depth, cell-free whole genome sequencing tracks thousands of personalized mutations in blood, identifying molecular relapse a median of 12.6-19 months before clinical progression and showing a stronger association with progression-free survival than standard bone-marrow tests (hazard ratio = 24 for BM-informed mutation lists). This scalable, comprehensive approach enables dynamic multiple myeloma monitoring and risk-adapted treatment.

**Summary Paragraph:** Minimal residual disease (MRD) monitoring is one of the strongest predictors of progression-free and overall survival in multiple myeloma (MM); as such, clinicians currently rely on invasive bone-marrow sampling that patients tolerate poorly for serial assessment. We show that longitudinal whole-genome sequencing of plasma cell-free DNA, guided by personalized mutation catalogs from diagnostic bone marrow, detects residual tumor DNA at ultra-low levels. This approach tracked disease dynamics across serial samples, achieved strong concordance with standard clinical MRD assays, accurately predicted progression-free survival, and detected subclonal evolution driving relapse. While developed initially using bone marrow reference, we found that cell-free DNA collected at baseline provided comparable mutational information to serve as a reference for subsequent MRD blood draws. This plasma-only classifier requires no marrow input, identified persistent disease when diagnostic marrow was insufficient for referencing, stratified relapse risk, and detected rising MRD probabilities a median of 19 months before progression. Together, these results establish cell-free DNA whole genome sequencing as a minimally invasive platform for comprehensive genomic surveillance in MM, reducing dependence on painful repeat marrow biopsies and enabling earlier, risk-adapted intervention. More broadly, this strategy can extend to diverse cancers to guide personalized therapy and identify relapse earlier.

## Introduction

Multiple myeloma (MM) is a hematological cancer characterized by clonal proliferation of malignant plasma cells in the bone marrow (BM).^1^ MM is a growing worldwide health threat, with prevalence more than tripling since 1990 due to global population aging and improved patient survival.^2^ Over the past decade, therapeutic advances such as proteasome inhibitors, immunomodulatory agents, and monoclonal antibodies have transformed multiple myeloma therapy, yielding unprecedented depth and durability of response.^3–6^ Despite these advances, myeloma remains a mostly incurable disease, and monitoring of minimal residual disease (MRD) has become an essential surrogate for progression-free survival (PFS) and overall survival (OS).^1,7^ Patients achieving and maintaining MRD negativity have significantly superior PFS and OS compared to those who do not.^7,8^ Reflecting this, in April 2024, the FDA’s Oncologic Drugs Advisory Committee endorsed MRD-negative complete response as a surrogate endpoint likely to predict PFS and OS, supporting its use in accelerated approval of new MM therapies.^9,10^

Standard MM MRD testing faces practical and biological constraints. Current approaches rely on BM aspirates analyzed by multiparameter flow cytometry (MFC), or targeted next-generation sequencing of immunoglobulin (Ig) gene rearrangements (NGS), both of which offer high sensitivity to detect malignant plasma cells (10^-5^ to 10^-6^).^11^ Serial BM sampling is invasive and painful, which limits its feasibility for longitudinal monitoring.^12,13^ Moreover, MM commonly exhibits patchy BM involvement, meaning that a single site biopsy may not be fully representative of the global tumor burden, thereby potentially leading to sampling errors.^14^ Extramedullary disease (EMD), present in approximately 8% of patients later in the disease course, is largely undetectable by BM-based assays.^9,11^ While positron emission tomography/computed tomography (PET/CT) is commonly used to detect extramedullary disease and assess metabolic tumor activity, its sensitivity for detecting MRD is limited without the use of a matched BM aspirate.^15,16^ Variability in specimen quality and hemodilution can further depress sensitivity and produce false-negative results.^11,17^ In addition, mass spectrometry (MS)–based assays of circulating monoclonal immunoglobulins have emerged as a minimally invasive MRD tool, with analytical sensitivity similar to MFC and Ig-NGS and feasibility for serial monitoring. However, MS measures only the intact protein product and provides no genomic or spatial information, limiting its capacity to capture clonal evolution or extramedullary disease.^18,19^

Cell-free DNA (cfDNA) analysis from peripheral blood (PB) presents a promising alternative that may better capture spatial heterogeneity, since tumor DNA is shed into the circulation from disparate disease sites.^20,21^ Prior studies show that myeloma-derived cfDNA recapitulates somatic alterations identified in BM. However, most MM cfDNA assays have employed targeted panels restricted to a small set of recurrently mutated genes or a single immunoglobulin rearrangement.^22–27^ Given that recurrent driver mutations in MM occur at low frequency and the mutational burden is modest relative to solid tumors, such narrow targeting increases the risk of false negatives and limits longitudinal tracking.^28,29^ In addition, assays that aim to detect rare mutant molecules at only a few hotspots or a single rearrangement often require large DNA inputs and very deep sequencing to reliably quantify MRD at very low levels.^30^

Whole genome sequencing (WGS) of cell-free DNA (cfWGS) provides a broader approach by simultaneously assessing thousands of genomic loci rather than a restricted gene panel or set of immunoglobulin loci, significantly increasing the likelihood that sequencing reads will capture tumor-derived mutations even at very low tumor fractions.^30–32^ This approach, exemplified by methods such as MRDetect, has achieved ultra-sensitive detection limits of approximately one tumor-derived molecule per 10^-5^ sequencing reads and has been successfully applied for MRD monitoring in breast cancer, B-cell lymphoma, and lung cancer.^30,32–34^ In addition, cfWGS enables fragmentomic readouts such as fragment size distributions and nucleosome footprints that are agnostic to any single mutation and can boost sensitivity at very low tumor fractions.^35^ Lastly, the non-invasive nature of blood sampling facilitates more frequent and readily available assessments, enabling near-real-time monitoring and dynamic evaluation of treatment response tailored to each patient’s clinical trajectory.

Despite these theoretical advantages, no study has validated cfWGS for MRD detection in MM, leaving its clinical utility uncertain. Here, we present a systematic evaluation of cfWGS for MRD detection in MM patients compared to standard BM-based assays, including MFC and NGS-Ig (clonoSEQ) across all stages of treatment, including induction, consolidation, maintenance, and relapse.

## Results

### Clinical Characteristics of Cohort

This study comprises cfWGS analysis of 163 blood cfDNA samples from 51 MM patients (median three samples per patient, range 1-7), collected over a median follow-up of 781 days (range 0– 2842). Baseline BM sequencing succeeded for 35 of 51 patients (69%), with most failures due to low CD138^+^ plasma-cell yield. Baseline cfDNA was available for 48 of 51 patients, all of whom were successfully sequenced (Fig. 1B). The analysis set was divided into a training cohort of 44 newly diagnosed, transplant-eligible patients treated with standard induction, most of whom proceeded to ASCT and lenalidomide maintenance at Canadian centers, and a test cohort of seven patients with heterogeneous treatment histories (non-standard induction, TCRT, or salvage therapy) to evaluate generalizability (Fig. 1A, Supplementary Table 1, **Methods**). Baseline characteristics are summarized in Table 1. ISS stages I/II/III comprised 44%/39%/17% of the training cohort and 0%/33%/67% of the test cohort. High-risk cytogenetics by FISH were observed in 25% of training patients and enriched in the test cohort at 60% of evaluable cases (Table 1).

**Figure 1:**
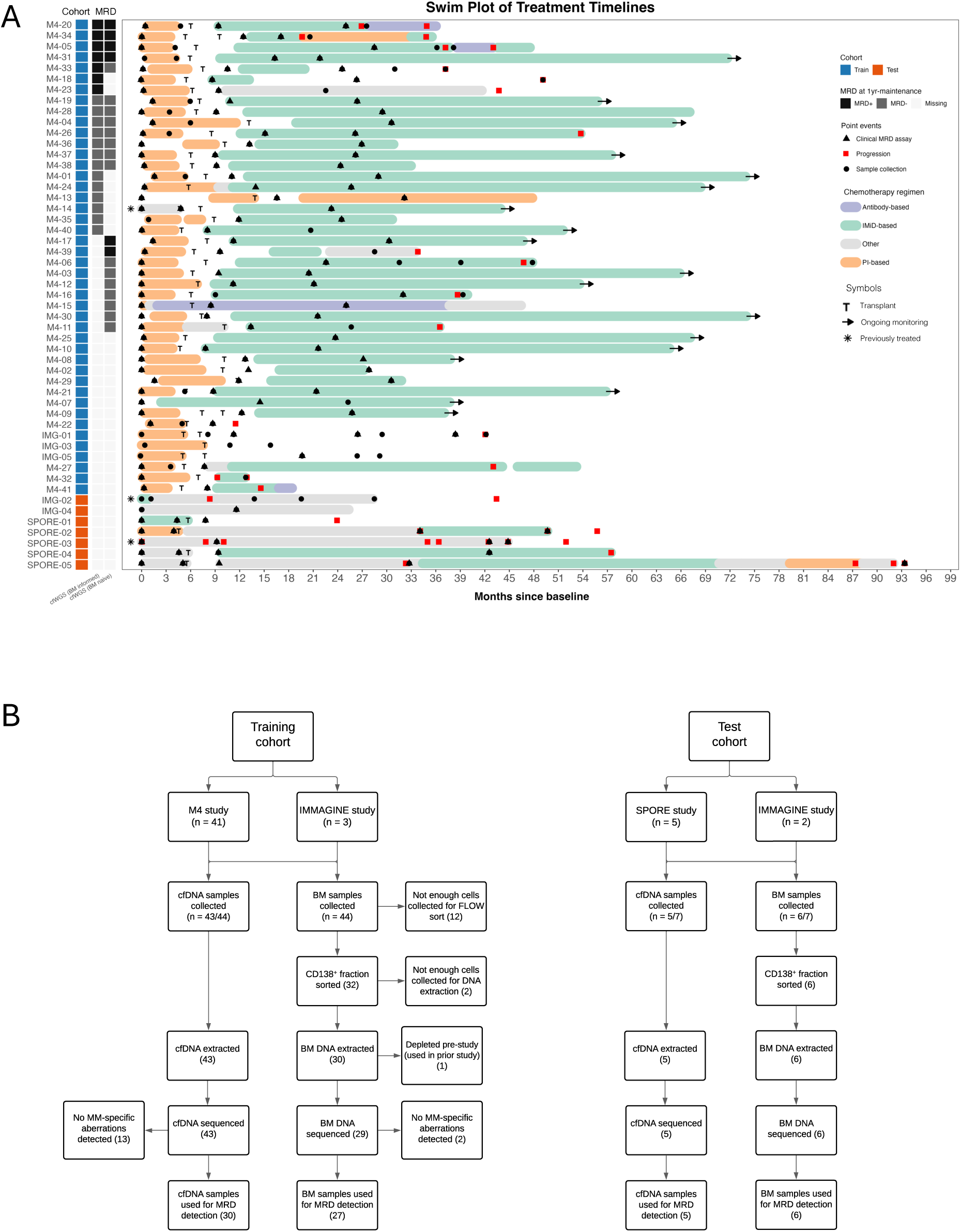
Overview of longitudinal treatment courses and baseline sample inclusion for MRD analysis in multiple myeloma. **(A)** Swim plot illustrating individual patient treatment timelines, beginning from diagnosis (day 0). Each horizontal bar represents an individual patient, grouped into training (blue) and test (orange) cohorts, and colored according to their chemotherapy regimen. Symbols denote key clinical and research events, including autologous stem-cell transplant (T), disease progression (red tick), and time points of bone-marrow (BM) or cell-free DNA (cfDNA) sampling used for cfWGS (black circles) and clinical MRD assays (black triangles). Arrows indicate ongoing follow-up, and asterisks mark patients previously treated before study enrollment either for smouldering myeloma (one patient) or for relapsed disease when the baseline sample corresponded to progression (two patients). The dot plot at left shows the change in circulating tumor DNA (ctDNA) cumulant variant allele fraction (cVAF) from baseline to the first on-treatment sample, derived from BM mutation lists when available, or from cfDNA when BM sequencing was unavailable at baseline. Positive values reflect an increase in ctDNA burden, while negative values indicate molecular response. **(B)** Flow diagram showing the collection, processing, and final inclusion of baseline bone marrow (BM) and cell-free DNA (cfDNA) samples for minimal residual disease (MRD) analysis in the training and test cohorts. The figure outlines sample availability and the quality control steps applied prior to tumor-informed MRD tracking using whole-genome sequencing. This overview highlights treatment diversity and cohort structure, facilitating the interpretation of longitudinal biomarker analyses.

**Table 1:** Clinical and Demographic Characteristics of Patients in the Training and Test Cohorts. Summary of baseline clinical and demographic characteristics for patients included in the training (N = 44) and test (N = 7) cohorts. Variables include gender, age group at diagnosis, International Staging System (ISS) stage, cytogenetic risk, myeloma immunoglobulin subtype, and ECOG performance status. Percentages are shown for each subgroup within the respective cohort. Missing values are reported where applicable.

| Variable | Training Cohort<br>N = 44<sup>1</sup> | Test Cohort<br>N = 7<sup>1</sup> | Total<sup>1</sup> |
| --- | --- | --- | --- |
| **Gender** |  |  |  |
| Female | 19 (43%) | 4 (57%) | 23 (45%) |
| Male | 25 (57%) | 3 (43%) | 28 (55%) |
| **Age Group** |  |  |  |
| <50 | 5 (11%) | 2 (29%) | 7 (14%) |
| 50-59 | 12 (27%) | 2 (29%) | 14 (27%) |
| 60-69 | 24 (55%) | 2 (29%) | 26 (51%) |
| 70-79 | 3 (6.8%) | 1 (14%) | 4 (7.8%) |
| **ISS Stage** |  |  |  |
| Stage I | 18 (44%) | 0 (0%) | 18 (41%) |
| Stage II | 16 (39%) | 1 (33%) | 17 (39%) |
| Stage III | 7 (17%) | 2 (67%) | 9 (20%) |
| (Missing) | 3 | 4 | 7 |
| **Cytogenetic Risk\*** |  |  |  |
| Standard Risk | 27 (75%) | 2 (40%) | 29 (71%) |
| High Risk | 9 (25%) | 3 (60%) | 12 (29%) |
| (Missing) | 8 | 2 | 10 |
| **Myeloma Ig Subtype** |  |  |  |
| IgA | 9 (21%) | 4 (67%) | 13 (27%) |
| IgG | 26 (62%) | 1 (17%) | 27 (56%) |
| Light Chain Only | 7 (17%) | 1 (17%) | 8 (17%) |
| (Missing) | 2 | 1 | 3 |
| **ECOG Performance Status** |  |  |  |
| 0 | 9 (43%) | 2 (33%) | 11 (41%) |
| 1 | 8 (38%) | 3 (50%) | 11 (41%) |
| 2 | 3 (14%) | 0 (0%) | 3 (11%) |
| 3 | 0 (0%) | 1 (17%) | 1 (3.7%) |
| 4 | 1 (4.8%) | 0 (0%) | 1 (3.7%) |
| (Missing) | 23 | 1 | 24 |

<sup>1</sup>n (%) \*High-risk cytogenetics defined as presence of t(4;14), t(14;16), t(14;20), del(17p), or amp(1q) by FISH.

### Baseline Genomic Characteristics of Cohort

To enable mutation-informed MRD tracking, we first evaluated the genomic features detectable by WGS in baseline samples (Extended Data Fig. 1). Disease-associated alterations were detectable in 94% of BM samples and 67% of cfDNA samples, defined by ≥1 somatic mutation, copy-number alteration (CNA), or chromosomal translocation (Extended Data Fig. 1). Plasma tumor fraction was modest (median ∼5.1% in the training and 5.3% in the test cohort), with ≥5% ctDNA in 51% and 60% of samples, respectively. Across matched BM–cfDNA pairs, concordance was high for translocations and variable for CNAs and mutations, increasing with higher ctDNA. Across all cohorts, we detected an average of 2,502 mutations per baseline BM (range: 106–8,721, SD = 1,616) and 2,498 per baseline cfDNA (range: 652–7,323, SD = 1,379) (Extended Data Fig. 2E), with cfDNA tumor fraction being the primary driver of cfDNA mutation recovery (Extended Data Fig. 2F). Detailed per-feature concordance, tumor-fraction-stratified sensitivity, FISH-vs-WGS probe-level analyses, and comparison of mutation counts between cohorts are provided in Supplementary Information, Extended Data Figs. 1–2 and Supplementary Tables 2-3. Collectively, these results highlight the feasibility of baseline genomic profiling from cfDNA and BM samples.

### Longitudinal cfDNA Dynamics During Therapy

In the training cohort (n = 44), cfDNA mutation metrics declined sharply during treatment, closely mirroring clinical response. Both BM- and cfDNA-derived mutation lists showed over two-order-of-magnitude reductions in the site-detection and cumulative VAF (cVAF) scores from diagnosis to first on-treatment time point (p < 0.001) (Fig. 2, Extended Data Fig. 3). These cfDNA-based measures correlated strongly with standard biomarkers, including M-protein, flow cytometry, and clonoSEQ (ρ ≥ 0.7, adjusted p < 0.05) (Extended Data Fig. 4). Fragmentomic features changed concordantly, reflecting a shift toward longer cfDNA fragments following treatment and lower tumor fraction, and were correlated with immunoglobulin-based disease burden and the proportion of tumor cells detected by MFC (adjusted p < 0.05). Additional quantitative comparisons, subgroup analyses, and biomarker correlations are provided in the Supplementary Notes. Together, these data demonstrate that cfDNA mutation features reliably capture therapy-aligned disease dynamics and support the development of cfWGS-based MRD classifiers (Extended Data Figs. 3-4, Supplementary Table 3).

**Figure 2:**
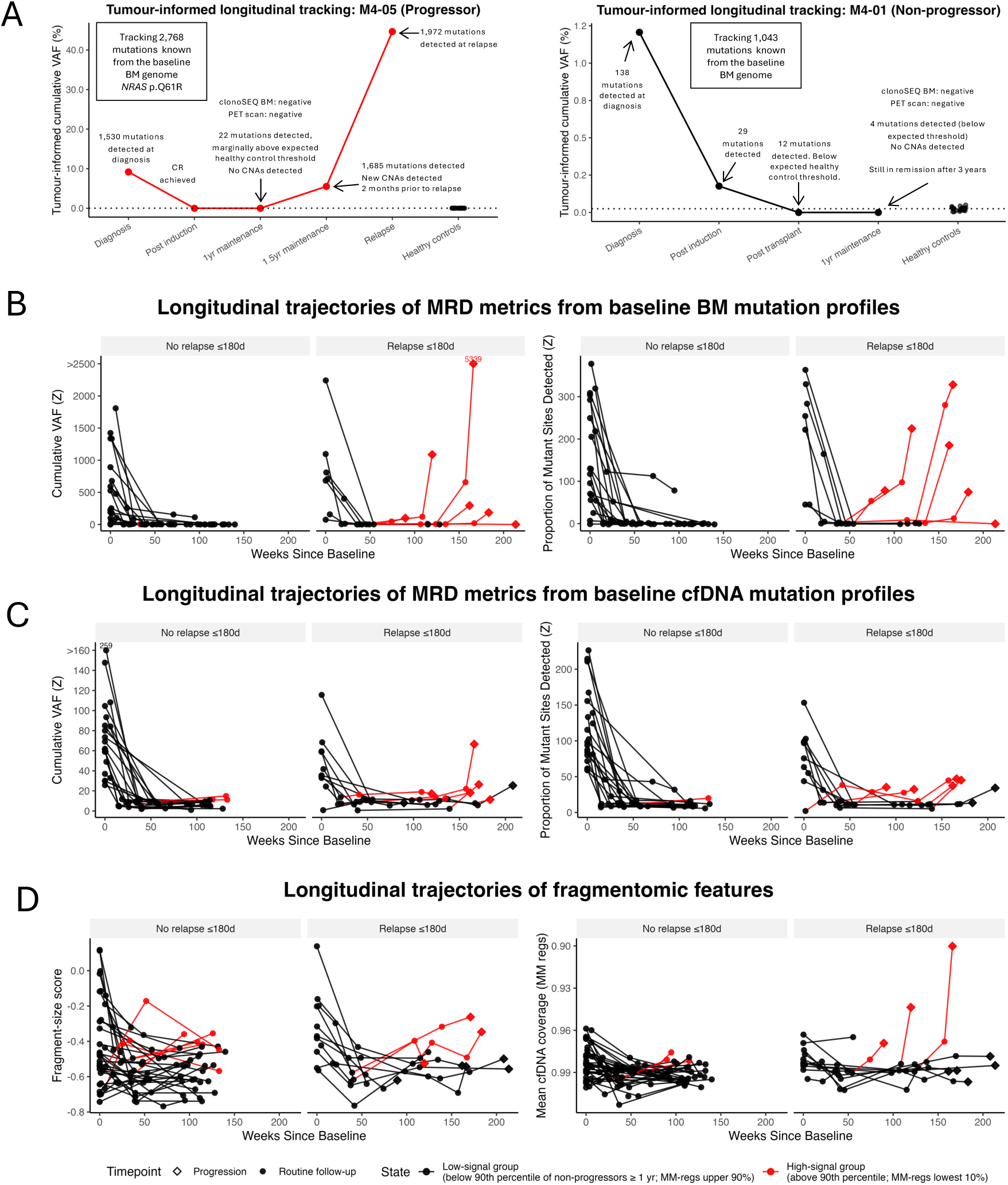
Longitudinal dynamics of tumor and peripheral blood-informed metrics for tracking minimal residual disease using cfDNA. **(A)** Representative examples of longitudinal, tumor-informed cumulative variant allele frequency (VAF) profiles from cfDNA analysis in one patient with clinical progression (left; patient M4-05, red line) and one patient without progression (right; patient M4-01, black line). Annotated points indicate clinical milestones, MRD assessment outcomes (clonoSEQ), and genomic observations (mutation burden, CNA detection). **(B-C)** Longitudinal trajectories of MRD metrics derived from bone marrow (BM) tumor mutation profiles (**B**) and peripheral blood (PB) cfDNA mutation profiles (**C**). Each line represents an individual patient. Panels show z-scores relative to healthy controls, computed from the cumulative VAF and the proportion of baseline-detected mutation sites. Patients are stratified by relapse within 180 days of the last available sample versus no relapse within that interval. Points are colored red when the sample value exceeded the 90th percentile of non-progressors after one year from the baseline timepoint. Samples collected at clinical progression timepoints are shown as diamonds. **(D)** Longitudinal trajectories of cfDNA fragmentomic features, including fragment-size score (left) and cfDNA coverage at active regulatory regions in multiple myeloma (right). Each line represents a patient. Points are colored red when exceeding the 90th percentile of non-progressors ≥1 year from baseline (for MM-regions coverage, lowest 10% as an inverse relationship is expected). Patients are stratified by relapse within 180 days of the last available sample versus no relapse. Collectively, these data demonstrate the potential of cfDNA-based molecular and fragmentomic analyses for sensitive, longitudinal MRD tracking and early relapse detection in multiple myeloma patients.

### Evaluating cfDNA Classifier Performance for MRD Detection

We next evaluated the performance of cfDNA-based MRD detection over time using the baseline mutation profiles identified in BM from patients who showed at least one disease-associated alteration at baseline and compared it against clinical MRD assays. In the training cohort, 42 samples from 26 patients were available with matched clinical MRD, collected over 4.8-38.5 months following diagnosis (median 16.8). We screened seven candidate cfWGS-based scoring rules, ranging from single-feature models to multi-feature combinations incorporating fragmentomic features (see **Methods**). Performance was assessed with nested 5 × 5 cross-validation. The top-performing model was the z-score of the proportion of sites detected relative to healthy controls (‘Sites Model’, see **Methods**), achieving a mean AUC of 0.86 ± 0.14 across the five outer folds (pooled AUC 0.81, 95% CI 0.67–0.96), with a mean sensitivity of 90%, specificity of 87%, accuracy of 88%, PPV of 87%, and NPV of 93% at the Youden threshold of 0.48 (Fig. 3A-B, Extended Data Fig. 5A, Supplementary Table 4). This model outperformed a Combined Model involving the cVAF z-score and Sites z-score (mean AUC = 0.821 ± 0.20), and the cVAF z-score only model (‘cVAF Model’) (AUC = 0.811 ± 0.17). Adding fragmentomic features did not improve performance (AUCs < 0.77; Extended Data Fig. 5A, Supplementary Table 4).

**Figure 3:**
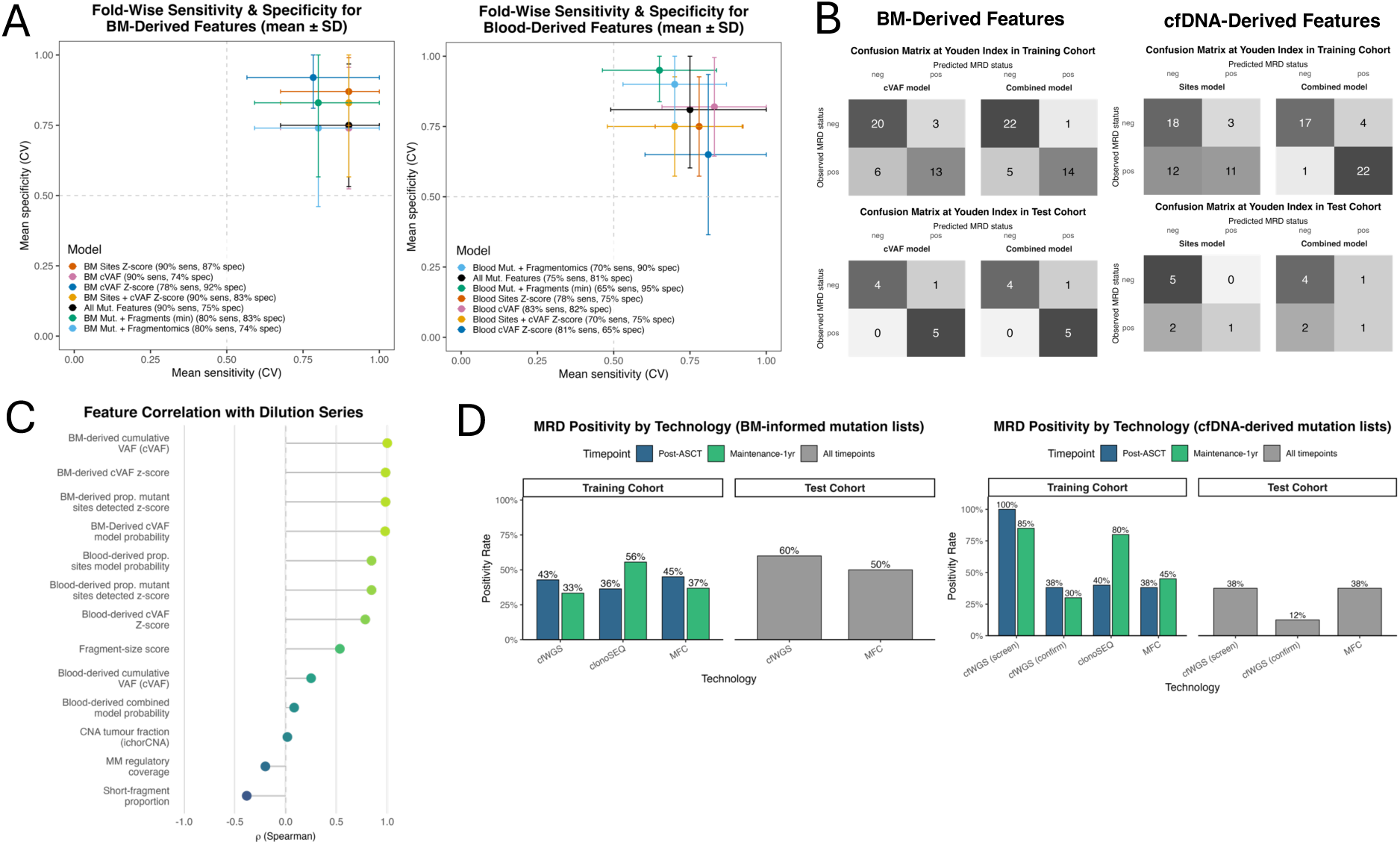
Analytical performance and concordance of cfDNA MRD classifiers. **(A)** Cross-validated mean sensitivity and specificity for each classifier, with error bars indicating ±1 standard deviation. Legend mapping: BM Sites Z-score = z-score of the proportion of baseline loci with ≥1 mutant read; BM cVAF = pooled mutant read fraction across baseline loci; BM cVAF Z-score = standardized cVAF; All Mut Features = Sites Z-score + cVAF + cVAF Z-score; BM + Fragmentomics = All Mut Features + fragment score (FS), mean GC-corrected coverage at MM regulatory sites, short-fragment proportion, and cfDNA tumor fraction (ichorCNA); BM + Fragments (min) = All Mut Features + FS + mean coverage only. Blood Sites Z-score: z-scored proportion of baseline loci with ≥1 mutant read; Blood cVAF: pooled mutant read fraction across baseline loci; Blood cVAF Z-score: standardized cVAF; All Mut Features: Sites Z-score + cVAF + cVAF Z-score; Blood + Fragmentomics: All Mut Features + fragment score (FS) + mean GC-corrected coverage at MM regulatory sites + short-fragment proportion + cfDNA tumor fraction (ichorCNA); Blood + Fragments (min): All Mut Features + FS + mean coverage only. **(B)** Confusion matrices displaying classifier performance at optimal Youden index thresholds for predicting MRD status in training (upper) and test (lower) cohorts, comparing models based on a combined model based on the cVAF z-score relative to healthy controls and proportion of sites detected z-score relative to healthy controls (“Combined Model”) and the cumulative variant allele frequency of all mutations (“cVAF model”). **(C)** Spearman’s rank correlations (ñ) between measured tumor fraction across a serial dilution series and individual cfDNA and BM-derived genomic and fragmentomic features used in MRD prediction. Features derived from BM and blood are shown separately, illustrating which metrics most strongly track tumor content at ultra-low fractions. **(D)** Comparison of MRD positivity rates determined by cfWGS versus clinical MRD assays (clonoSEQ and multiparametric flow cytometry [MFC]) across post-autologous stem-cell transplant (ASCT) and maintenance therapy time points in both training and test cohorts. Together, these panels establish analytical validation and clinical concordance of cfDNA MRD classifiers with clinical MRD assays, supporting their validity as reliable tools for non-invasive disease monitoring.

To assess performance using all available data, we refitted each model on the full training cohort and evaluated performance at the Youden threshold identified during cross-validation. The refit combined model performed best with 74% sensitivity and 96% specificity, with an overall accuracy of 84% (NPV = 82%, PPV = 93%) (Fig. 3B, Extended Data Fig. 5B, Supplementary Table 5). When classifiers were compared at a fixed 95% sensitivity, mimicking a high-surveillance use case, the Combined Model maintained 13% specificity (accuracy = 56%), which was lower than the cVAF and Sites Models (17% specificity, 58% accuracy) (Extended Data Fig. 5B, Supplementary Table 5). At the Youden threshold, the Combined Model balances sensitivity and specificity; under high-surveillance settings, it preserves sensitivity with expected specificity trade-offs.

Lastly, we evaluated these models on 10 samples from 5/7 patients in the test set, which had matched clinical MRD results (all MFC [10^-5^ sensitivity], collected over 3.9-49.7 months following diagnosis, median 11 months). In this set, the combined model achieved 100% sensitivity and 80% specificity (accuracy = 90%), while the cVAF Model achieved 100% sensitivity at 80% specificity (accuracy = 90%), and Sites Model achieved 80% sensitivity and 80% specificity (accuracy = 80%) (Fig. 3B, Extended Data Fig. 5C, Supplementary Table 6). Despite the small test set, these point estimates support the generalizability of BM mutation-based mutation models to independent samples.

### Quantifying Sensitivity and Limit of Detection

To determine the limit of detection of our cfDNA-based classifiers, we generated a dilution series by mixing baseline-positive and MRD-negative cfDNA samples at known tumor fractions (see **Methods**). We then assessed how well each candidate MRD metric correlated with the known tumor fraction across this series. Spearman correlation revealed that the cVAF demonstrated a perfect monotonic relationship with the known tumor fraction (ρ = 1.00, p < 0.001), followed closely by the z-score of the cVAF relative to healthy controls (ρ = 0.98, p < 0.001). The z-score of the proportion of sites detected performed equivalently (ρ = 0.98, p < 0.001) (Fig. 3C, Extended Data Fig. 5D). Other fragmentomic and sequencing coverage metrics exhibited only modest or negligible correlations (ρ ≤ 0.53) (Fig. 3C, Supplementary Table 7).

Using the optimal classification threshold identified by the Youden index during cross-validation, the Combined Model classified all samples with ≥0.061% tumor fraction as MRD-positive, whereas the cVAF Model did so for all samples with ≥0.011% tumor fraction (Extended Data Fig. 5D, Supplementary Table 7). Prioritizing maximal MRD sensitivity and the lowest empirical limit-of-detection, we selected the cVAF Model at its optimal threshold as our training classifier. These results demonstrate that cfWGS reliably tracks tumor content and establish an empirical limit of detection at 0.011% VAF, defining the range where cfWGS is informative and where an ultrasensitive orthogonal assay may be required.

### Concordance with Clinical MRD Assays

To contextualize cfWGS performance alongside MFC and the clonoSEQ NGS-based MRD assay, we compared positivity and concordance at landmark time points using all samples with at least one clinical MRD result available. This included 20 of 21 post-ASCT samples and 19 of 21 samples at 1-year maintenance. At the post-ASCT time point, cfWGS detected MRD positivity in 43% of patients (9/21), comparable to MFC (45%; 9/20) and clonoSEQ (36%; 4/11) (Fig. 3D). At 1-year maintenance, cfWGS was positive in 33% of patients (7/21), similar to MFC (37%; 7/19) and lower than clonoSEQ (56%; 5/9) (Fig. 3D). Of the 8 patients who were negative at both timepoints by MFC, 7 (87.5%) were also negative by cfWGS. At post-ASCT and 1-year maintenance, cfWGS positivity is broadly comparable to MFC and clonoSEQ.

To further assess assay performance, we examined concordance between cfWGS and clinical MRD tests at each time point. At post-ASCT, cfWGS demonstrated high concordance with both clinical assays, agreeing with clonoSEQ in 9 of 11 samples (82%, PPV = 75%, NPV = 86%) and with MFC in 16 of 20 samples (80%, PPV = 78%, NPV = 82%) (Extended Data Fig. 5E). All discordant results occurred near or below the empirical limit of detection of one or more assays (Fig. 4A). For example, in patient M4-13 (17 months post-diagnosis), clonoSEQ detected a low-level clone (VAF = 0.013%) that was not identified by cfWGS or MFC, suggesting residual disease below the cfWGS detection threshold of 0.011% VAF in plasma. Conversely, patient M4-33 (11 months post-diagnosis) was cfWGS-positive while clonoSEQ was negative at the clinically reported LOD of 1.23 per million cells. Nevertheless, clonoSEQ detected the tumor clonotype at 0.99 per million cells, below the clinical reporting threshold due to limited cell quantity and quality in the bone marrow sample. The patient subsequently progressed two years later, consistent with possible sampling bias or patchy marrow involvement. In contrast, several MFC-positive but cfWGS-negative patients (e.g., M4-20 and M4-40) had extremely low marrow plasma cell percentages (<0.006%), suggesting a localized disease burden undetectable in plasma (Fig. 4A, Supplementary Table 8).

**Figure 4:**
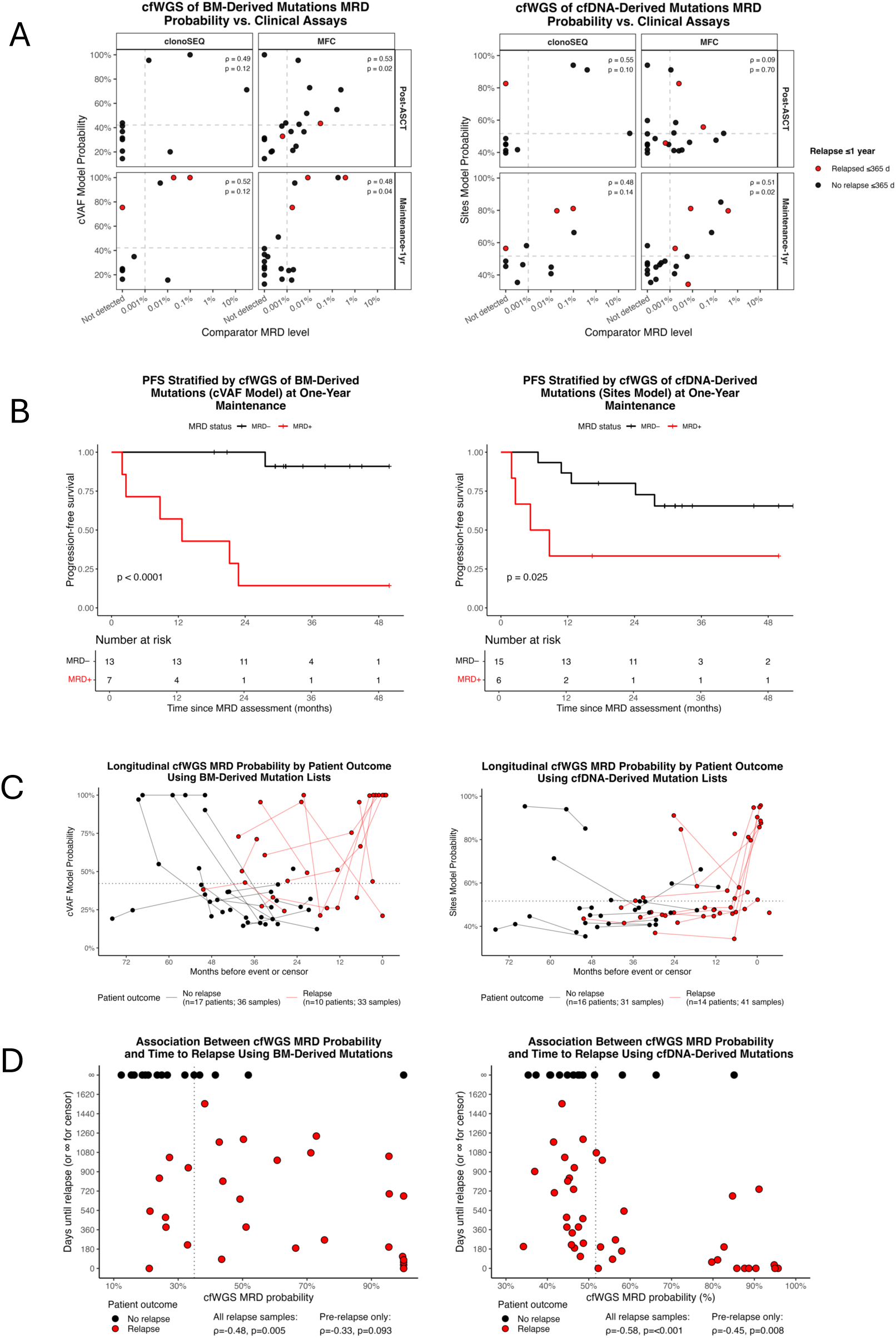
cfDNA MRD tracking captures relapse risk and survival dynamics. **(A)** Comparison of cfWGS-based MRD probability with clinical MRD assays in the training cohort. Each point represents one sample, showing cfWGS-derived MRD positive probability (y-axis) versus clinically established MRD status (x-axis). Detection patterns illustrate concordance and discordance among measurement approaches, stratified by clinical outcome (IMWG-defined progression vs. no progression within one year of sampling). For BM–derived features, MRD probability was estimated using the cVAF Model, based on the cVAF z-score relative to healthy controls. For blood-derived features, the Sites Model, which uses the z-score of the proportion of detected mutant sites relative to healthy controls, was applied. **(B)** Kaplan–Meier curves showing time to progression stratified by cfWGS-based MRD status in a landmark analysis at one year of maintenance therapy, underscoring the prognostic relevance of cfDNA-based MRD detection. Numbers at risk are shown below each time point. **(C)** Longitudinal cfWGS MRD probability trajectories for individual patients. Each line tracks one patient’s MRD probability (%) using the cVAF Model (for BM-derived mutation lists) and Sites Model (for blood-derived mutation lists) over months before relapse or censor; blue = no relapse, green = relapse. The horizontal dotted line marks the Youden threshold used to determine a positive MRD call. **(D)** Scatterplot showing the relationship between cfWGS-derived MRD probability and time to relapse or censoring. Each point represents a sample, with relapse events in red and censored cases (who have not progressed) in black. For patients who relapse (red), all samples following diagnosis are included, whereas for patients who remain in remission (black), their latest available treatment timepoint is included (typically during maintenance). Higher MRD probabilities are associated with shorter time to relapse, suggesting the predictive value of cfDNA for impending progression. Collectively, these analyses demonstrate that cfWGS-derived MRD probabilities align with clinical assays, stratify relapse risk in landmark analyses, capture longitudinal disease dynamics, and predict time to progression, highlighting the clinical utility of cfDNA for non-invasive monitoring and early relapse detection.

At 1-year maintenance, concordance patterns were similar. cfWGS and clonoSEQ agreed in 6 of 9 cases (67%) (PPV = 75%, NPV = 60%), while cfWGS and MFC were concordant in 15 of 18 cases (83%) (PPV = 83%, NPV = 83%) (Extended Data Fig. 5F). As at earlier time points, discordances clustered near assay detection limits (Fig. 3G). Patients M4-19 and M4-28 (26 and 29 months post-diagnosis, respectively) were clonoSEQ-positive with a VAF in BM of <0.010%, below the cfWGS detection threshold. In contrast, patient M4-05 (28 months post-diagnosis) was cfWGS-positive (cVAF 0.08%) while clonoSEQ reported no detectable Ig fragments (specimen-specific LOD of 10⁻⁵). MFC confirmed low-level residual plasma cells in M4-05 (0.0017%), and this patient progressed within 9 months. Similarly, other cfWGS-negative, MFC-positive cases (e.g., M4-01) again exhibited extremely low BM plasma cell content (<0.001%), reinforcing the sensitivity boundaries and complementary detection profiles of each modality (Fig. 4A, Supplementary Table 8). Most discordances arose close to each assay’s limit of detection and reflected compartmental sampling (blood vs focal marrow), indicating complementary profiles.

We next evaluated the performance of our model in the test cohort. cfWGS identified MRD positivity in 60% of cases (6/10), compared to 50% by MFC (5/10 with matched cfWGS), yielding 100% sensitivity, 80% specificity, 83% PPV, and 100% NPV when MFC was used as the comparator (Fig. 3D, Extended Data Fig. 5G). Overall agreement between cfWGS and MFC was 9/10 (90%) (Extended Data Fig. 5G, Supplementary Table 8). The sole discordant case was positive by cfWGS but negative by MFC; notably, this patient experienced biochemical progression within 30 days of the MFC and cfDNA assessment. This discrepancy likely reflects spatial heterogeneity or sampling bias in the bone marrow aspirate, leading to false-negative results by MFC. In the test cohort, cfWGS closely mirrored MFC and additionally identified an MFC-negative case that soon progressed, underscoring its potential utility for early relapse detection.

Taken together, these findings demonstrate that cfWGS yields comparable MRD positivity rates to both clonoSEQ and MFC across training and test cohorts, with high concordance at clinically relevant time points.

### Detection of Progression

After a median follow-up of 48.9 months (IQR 38-59; range 9-79), 16 of 44 patients (36%) in the training cohort experienced disease progression (Fig. 1A). We next evaluated the predictive performance of each MRD modality in those patients with cfWGS data at any landmark time points, enabling head-to-head comparison across assays. At the post-ASCT time point, cfWGS was MRD positive among 67% (6/9) of eventual progressors, while MFC detected 75% (6/8) and clonoSEQ 67% (2/3) (Extended Data Fig. 6A). By the one-year maintenance time point, sensitivities were similar across modalities; cfWGS detected 86% (6/7), clonoSEQ 75% (3/4), and MFC 67% (4/6) of patients who later progressed (Extended Data Fig. 6A). After ASCT and at 1-year maintenance, cfWGS identifies a similar fraction of future relapses as MFC/clonoSEQ, suggesting comparable prognostic utility.

MRD status at 1-year maintenance stratified relapse risk. We selected the 12-month on-maintenance landmark, consistent with FDA ODAC recommendations recognizing MRD at 9- or 12-months post-maintenance initiation as an appropriate regulatory endpoint.^10^ Patients who were MRD-negative by cfWGS had 100% relapse-free probability at 24 months after the 1-year maintenance MRD test, compared to 14% for MRD-positive patients (HR = 24.14; 95% CI 2.75– 211.73; Fig. 4B, Extended Data Fig. 6B). Median time to progression from the MRD test was not reached in the MRD-negative group, whereas MRD-positive cases had a median of 12.6 months. MFC showed a similar pattern (n = 20 evaluable patients), with 91% relapse-free probability at 24 months for MRD-negative patients versus 43% for MRD-positive (HR = 4.28; 95% CI 0.78–23.57) (Extended Data Fig. 6B-C). clonoSEQ data were available for a small subset of patients at this time point (n=11) and showed similar stratification to MFC (HR = 3.07; 95% CI 0.32–29.76) (Extended Data Fig. 6B, D). With only seven events among 21 evaluable patients, statistical power was limited (power = 14% for HR = 2.0, minimum detectable HR for 80% power is 84.8). The association observed for cfWGS (HR = 24.14) was statistically significant, but the wide confidence interval reflects uncertainty, suggesting these results are descriptive and hypothesis-generating, warranting validation in larger cohorts. Accordingly, Kaplan-Meier curves and univariable Cox models are reported descriptively without emphasis on log-rank p-values (Fig. 4C). Notably, MRD status at the earlier post-ASCT time point was less discriminative (HR = 3.2; 95% CI 0.8–12.86), likely reflecting fewer early relapse events (Extended Data Fig. 6B, E-F). Despite limited power, cfWGS MRD negativity at 1-year maintenance associates with favorable relapse free survival probability and merits validation in larger cohorts.

To explore the temporal dynamics of relapse detection, we examined 27 cfWGS samples from 10 patients in the training cohort collected after induction therapy, at post-ASCT, or during maintenance, prior to clinical progression (median 645 days before progression; IQR 811 days; range 32-1,533). The lowest MRD positive probability occurred a median of 729 days before progression (IQR 476 days; range 85-1,232). From that lowest point, the first increase in MRD-positive probability occurred a median of 427 days later (IQR 374 days; range 91-742), which was a median of 384 days before IMWG-defined clinical progression (IQR 368 days; range 58-1,076) (Fig. 4C). Across samples, there was an inverse association between cfWGS MRD probability and days until relapse (Spearman’s ρ = -0.33, p = 0.09; ρ = -0.48, p = 0.005 when including samples at the time of relapse, n = 33), indicating that higher MRD probabilities tend to occur closer to the date of clinical progression (Fig. 4D). Rising cfWGS MRD probabilities preceded clinical relapse by a median of 12.6 months, underscoring their potential for early molecular-resurgence monitoring.

In the test cohort, sampling times were heterogeneous, so we assessed each assay’s ability to predict progression within fixed time windows (180 and 365 days). Among cfWGS samples, five from two patients relapsed within 180 days and seven from four patients relapsed within 365 days. cfWGS identified 100% of 180-day relapses (5/5; specificity 58%) and 86% at 365 days (6/7; specificity 60%). MFC detected 80% of 180-day relapses (4/5; specificity 57%) and 67% at 365 days (4/6; specificity 50%) (Extended Data Fig. 6G; Supplementary Table 9). These results suggest that cfWGS performs comparably to MFC in predicting near-term relapse. However, modest predictive power and limited sample size underscore the need for larger, prospective studies to validate these observations. Windowed analyses indicate near-term relapse prediction comparable to MFC, though estimates remain imprecise due to small sample size.

### Testing cfDNA-Derived Baseline Classifiers When BM Is Unavailable

To ensure cfDNA-based MRD surveillance is feasible even when diagnostic BM-derived DNA cannot be obtained, such as in cases of low CD138⁺ cell yield, which occurred in 14/44 patients (32%) in the training cohort (Fig. 1B), we developed classifiers using mutations identified directly from baseline peripheral blood cfDNA.

#### Model Training and Performance

We initially trained and assessed classifiers using 52 MRD samples from 25 patients in the training cohort who had detectable disease-associated features at diagnosis (see Methods). Among these, 44 MRD samples from 28 patients had matched clinical MRD results and were included for model training, collected over 7.6-38.5 months following diagnosis (median 16.7). Importantly, this subset contained 16 samples from 11 patients whose diagnostic BM samples either yielded insufficient cells (n = 10 patients) or failed sequencing QC (n = 1) (Fig. 1B). We evaluated seven distinct cfDNA-based models using different molecular and fragmentomic features (see **Methods**). The best-performing classifier in the training set was a Combined Model using all blood-derived mutation and fragmentomic features (the cVAF, z-score of the cVAF relative to healthy controls, z-score of the proportion of sites detected relative to healthy controls, fragment size score, proportion of short fragments, coverage at MM-specific sites, and ichorCNA tumor fraction, see **Methods**), achieving a mean AUC of 0.79 ± 0.09 across the five outer folds (pooled AUC 0.741, 95% CI 0.59–0.89), with a mean sensitivity of 70%, specificity of 90%, accuracy of 79%, PPV of 91%, and NPV of 74% at the Youden threshold of 0.472 (Fig. 3A-B, Extended Data Fig. 7A, Supplementary Table 4). This outperformed the cVAF Model (mean AUC = 0.78 ± 0.12), proportion of sites detected z-score model (‘Sites Model’, mean AUC = 0.69 ± 0.07), and other tested models (Fig. 3A-B, Extended Data Fig. 7A-B, Supplementary Table 4).

In the small test cohort (8 samples from 4 of 7 patients with matched clinical MRD data, collected 4.9-93.4 months after diagnosis; median follow-up 11 months), the Combined Model achieved 63% accuracy, with moderate sensitivity (67%) but low specificity (20%). The Sites Model preserved specificity and achieved higher overall accuracy (75%; sensitivity 33%, specificity 100%), albeit with low sensitivity (Fig. 3B, Extended Data Fig. 7C, Supplementary Table 6). These results should be interpreted with caution, given the small sample size and non-independence of observations. Together, these results suggest that while a multi-feature cfDNA model performed best in training, a simpler sites-z-score classifier generalized more consistently in the small test set, motivating its use as the primary model with conservative interpretation.

#### Quantifying Sensitivity and Limit of Detection

To empirically define cfDNA-based MRD detection thresholds, we repeated a dilution-series experiment spanning tumor fractions from 0.009% to 0.688% (n = 9 samples). The Sites Model MRD+ probability correlated strongly with tumor fraction (Spearman’s ρ = 0.85, p <0.01), surpassing other tested metrics (Fig. 4C). Fragmentomic features and the combined model probability had minimal correlation with tumor fraction (ρ ≤ 0.1; Fig. 4C, Extended Data Fig. 7D, Supplementary Table 7). However, applying the initial Youden-derived probability threshold from the Sites Model failed to detect even the highest dilution (0.688%; Fig. 4D). To improve detection of low-level disease, we introduced a lower “screening” threshold (0.380), successfully identifying tumor fractions as low as 0.061% (sensitivity 91%, specificity 5% in training cohort). The original threshold (0.517) was retained as a confirmatory cutoff, maintaining higher specificity (86%) at reduced sensitivity (48%; Extended Data Fig. 7E). Thus, samples falling between these two thresholds represent a “gray zone,” necessitating repeat or orthogonal confirmation. Although the Sites Model exhibited lower classification performance in the training cohort compared to the combined model, we selected it as the primary MRD classifier due to its strong correlation with tumor fraction in controlled dilution-series experiments and performance in the independent test set. These findings suggest improved generalizability and enhanced sensitivity to low-level disease.

#### Concordance with Clinical MRD Assays

We assessed concordance between cfDNA-based MRD (using the Sites Model) and clinical assays at post-ASCT and 1-year maintenance in samples with at least one clinical MRD result (n = 21/23 and 20/23, respectively). At post-ASCT, the cfDNA-based screen threshold identified 100% (21/21) as MRD-positive, whereas the confirmatory threshold was positive in 38% (8/21), closer to clinical assays: 40% (4/10) by clonoSEQ and 38% (8/21) by MFC (Fig. 3D). At 1-year maintenance, screening positivity was again high (85%; 17/20), while confirmatory positivity was more conservative (30%; 6/20), lower than MFC (35%; 9/20) and clonoSEQ (80%; 8/10; Fig. 3D). Of the 8 patients that were negative at both timepoints by MFC, 7 (87.5%) were also negative by cfWGS. Due to excessive positivity using the screening threshold, subsequent analyses used the confirmatory threshold.

At post-ASCT, confirmatory cfDNA-based MRD demonstrated strong concordance with clonoSEQ (8/10, 80%; PPV 75%, NPV 83%) and MFC (15/21, 71%; PPV 62%, NPV 77%; Extended Data Fig. 7F). Discordances were predominantly cfDNA-negative cases with intermediate (“gray-zone”) probabilities, positive by clinical MRD. Notably, patient M4-39 (10 months post-diagnosis) was cfDNA-positive but MFC-negative and progressed just over two years later, whereas patient M4-41 (8 months post-diagnosis) was cfDNA-positive but clonoSEQ-negative and progressed within one year (Fig. 4A, Supplementary Table 8). Interestingly, all four false negatives were correctly identified when fragmentomic features were incorporated using the Combined Model, which achieved high concordance with clinical assays (agreement with MFC: 86%, PPV 73%, NPV 100%; agreement with clonoSEQ: 90%, PPV 80%, NPV 100%; Supplementary Table 10).

At 1-year maintenance, cfDNA-based MRD showed 44% concordance (4/9) with clonoSEQ (PPV 75%, NPV 20%) and 79% (15/19) with MFC (PPV 100%, NPV 71%; Extended Data Fig. 7G). Again, discordances clustered near assay detection thresholds, with cfDNA-negative discordances exhibiting extremely low disease burden by clinical assays (<0.002% by MFC, <0.01% by clonoSEQ; Fig. 4A; Supplementary Table 8). Notably, 5 of 6 patients negative by cfWGS but positive by clinical testing were reclassified as positive when incorporating fragmentomic features using the Combined Model, which showed stronger concordance with clinical assays (MFC: 84%; PPV 75%, NPV 100%; clonoSEQ: 78%; PPV 86%, NPV 50%; Supplementary Table 10). Additionally, patient M4-05 was cfDNA-positive but clonoSEQ-negative at maintenance (28 months post diagnosis) yet progressed within a year, suggesting residual disease missed by immunoglobulin-based methods (Supplementary Table 8). Discrepancies concentrated near the assays’ limits of detection, with cfDNA-negative misclassifications occurring at very low disease burden but recoverable by incorporating fragmentomic features.

In the test cohort (8 samples, 4 patients), cfDNA-based MRD was concordant in 6/8 cases, missing two cases with 0.9% and 0.1% cells in BM by MFC; however, the case with 0.9% cells was captured using the Combined Model (Extended Data Fig. 7H, Supplementary Tables 8-9). Overall, these findings support the utility of blood-derived mutation classifiers for MRD monitoring when baseline BM samples are unavailable or insufficient. Collectively, these data indicate that the confirmatory cfDNA classifier aligns more closely with clinical assays than the screening threshold and, in select discordant cases, can flag residual disease missed by marrow-based testing. This supports its use for MRD monitoring when baseline BM material is unavailable or yields insufficient material for mutation list generation.

#### Detection of Progression

We evaluated the capacity of the blood-based cfDNA model to predict relapse. At post-ASCT, cfWGS using the Sites Model was MRD positive in 46% of patients who eventually progressed (5/11) and 73% (8/11) when incorporating fragmentomic features using the Combined Model, versus 56% (5/9) by MFC and 60% (3/5) by clonoSEQ (Extended Data Fig. 8A). By 1-year maintenance, cfWGS using the Sites Model detected 45% (4/9) of cases and 78% (7/9) with the Combined Model, compared to 71% by MFC (5/7) and 75% by clonoSEQ (75%; 3/4) (Extended Data Fig. 8A). At 1-year maintenance, cfDNA-negative patients showed markedly better relapse-free probability (80% vs 33% at 24 months; HR 4.18; 95% CI 1.08–16.16 using the Sites Model; and HR = 2.38; 95% CI 0.49–11.5 using the Combined Model), which stratified patients comparably to MFC (90% vs 56%; HR 4.23; 95% CI 0.81-22.1) (Fig. 4H, Extended Data Fig. 8B-C). Due to limited events, the analyses remained underpowered; thus, these findings are descriptive and hypothesis-generating. In the test cohort, blood-based cfDNA MRD using the Sites Model detected 50% of patients progressing within 180 days (2/4; specificity 50%) and 50% within 365 days (3/6; specificity 50%), compared to 75% (3/4; specificity 57%) at 180 days by MFC and 60% (3/5; specificity 50%) at 365 days (Extended Data Fig. 8D; Supplementary Table 10). Taken together, these estimates, while imprecise given few events, suggest that cfWGS provides relapse risk stratification at 1-year maintenance comparable to clinical MRD assays, with similar sensitivity in the test cohort but limited specificity.

Lastly, longitudinal cfDNA monitoring of 33 samples from 13 patients collected after induction, post-transplant, or during maintenance revealed that increases in MRD probability were typically detectable well in advance of IMWG-defined clinical progression, with the first rise observed a median of 578 days before progression (IQR 499 days; range 79-1076 days) (Fig. 4C). Furthermore, cfWGS MRD probability was significantly inversely correlated with time to relapse (Spearman’s ρ = -0.45, p = 0.008), indicating that higher MRD probabilities tend to occur closer to the time of progression (Fig. 4D). Overall, rising cfWGS MRD probabilities typically precede clinical progression by a median lead time of 19 months, supporting their potential for early molecular-resurgence monitoring.

### Exploratory Analysis of Fragmentation-Based MRD Detection Without a Baseline Sample

We next evaluated our ability to classify MRD status without baseline tumor sequencing, relying solely on cfDNA fragmentation features. This analysis expanded our evaluable dataset to include samples without high-quality baseline BM or cfDNA, yielding a total of 65 samples from 41 patients in the training cohort with matched clinical MRD outcomes. We assessed six models incorporating individual or combined fragmentomic features using 5×5 nested cross-validation (**Methods**). The Coverage-at-MM-sites Model, leveraging the mean coverage at MM-specific chromatin accessibility regions, showed the highest discrimination, albeit modest (mean AUC = 0.63 ± 0.05 across the outer folds, pooled AUC of 0.56, 95% CI 0.42–0.70). This model was followed by a model based on the proportion of short cfDNA fragments (mean AUC = 0.63 ± 0.11). Models using other features or feature combinations showed poorer performance (Supplementary Table 4, Extended Data Fig. 9A). The Coverage-at-MM-sites Model achieved a mean sensitivity of 80% and specificity of 60% (accuracy = 56%) for MRD detection at the Youden index threshold (Extended Data Fig. 9B). This coverage model represented the best accuracy among fragmentomics-only models tested, with others achieving only 31–54% mean accuracy across outer folds, showing comparable performance when refit and evaluated on the full cohort using the Youden-optimized threshold (Supplementary Tables 4-5, Extended Data Fig. 9C, E).

Twelve samples from six patients were available in the test cohort. Using training-derived thresholds, a Combined Model using the fragment score and Coverage-at-MM-sites achieved 90% accuracy in the test set (sensitivity 100%, specificity 80%). In contrast, the performance using Coverage-at-MM-sites alone achieved 58% accuracy (Extended Data Fig. 9D, F, Supplementary Table 6). Collectively, these results indicate that fragmentomics alone provides only modest discrimination and inconsistent performance in the test cohort, supporting the use of mutation-informed cfDNA models and positioning fragmentation features as complementary rather than standalone MRD markers in MM. Although previous studies have reported that certain fragmentomic features can contribute to cancer detection,^36^ the specific metrics evaluated here did not yield sufficient standalone performance.

### Evaluating Subclonal Evolution using cfDNA

Given the clinical importance of identifying therapy-resistant subclones before overt progression, we profiled CNAs longitudinally by cfWGS. To investigate whether cfDNA could capture the evolution of high-risk subclones over time, we analyzed 40 cfDNA samples from 10 patients with paired diagnosis and progression time points, with a median of 4 samples per patient (range 2–5), spanning both cohorts. In 5 of 10 patients (50%), we observed subclonal evolution marked by the *de novo* acquisition of MM-specific CNAs at progression, including amp(1q), del(13q), and del(17p), of which 4/5 patients experience a high-risk CNA. For example, patient M4-05 developed an amp(1q) in cfDNA 32 days before biochemical progression, which became more prominent at the progression visit (Fig. 5A). Patient SPORE-0005, who had no detectable CNAs at diagnosis, acquired an amp(1q) event at relapse, detectable in cfDNA. Similarly, patients M4-06, M4-16, and M4-32 showed emergent high-risk CNAs at progression, including concurrent amp(1q), del(13q), and del(17p) events (Fig. 5B).

**Figure 5:**
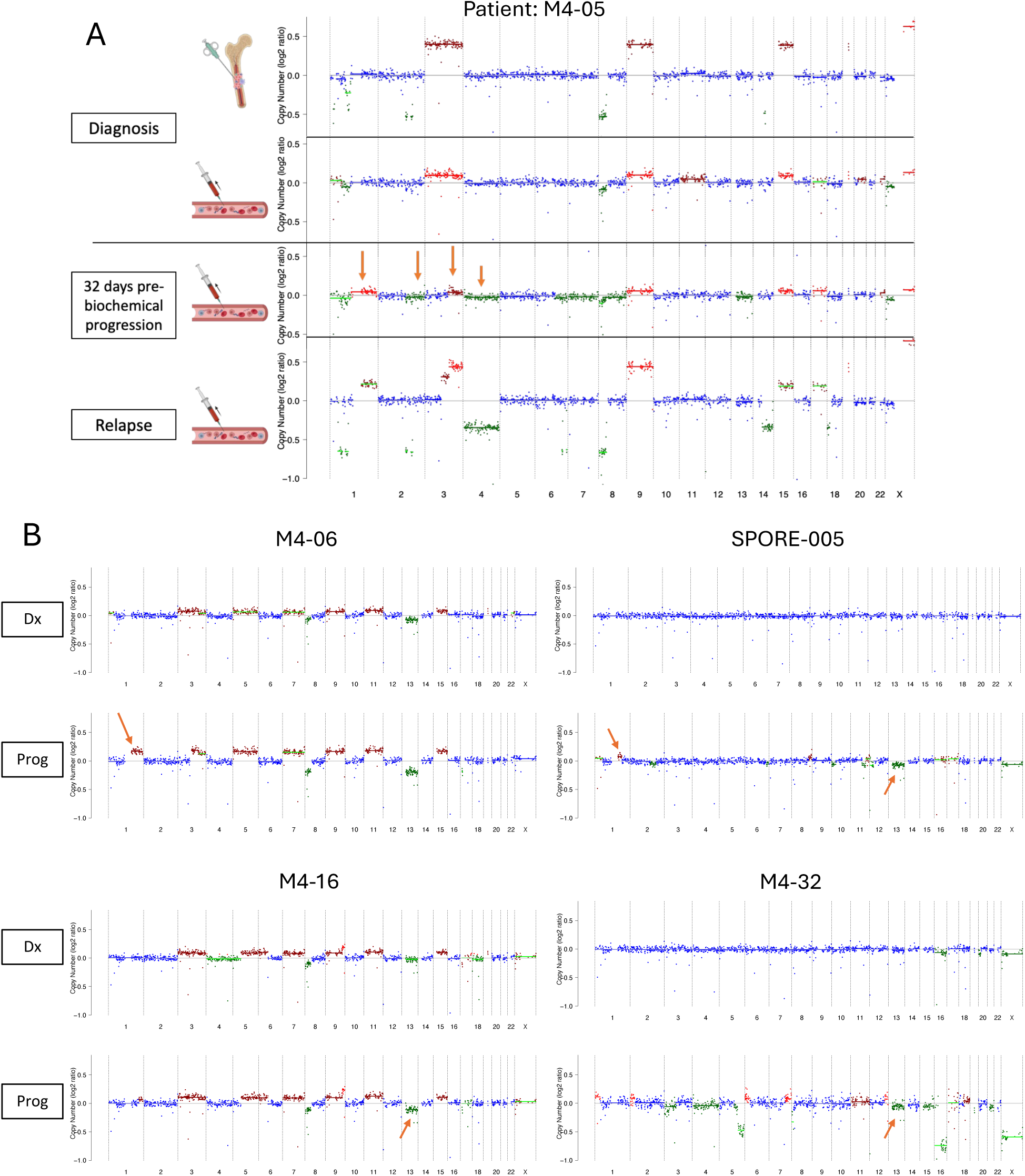
Longitudinal detection of emerging copy-number subclones in cfDNA. **(A)** Longitudinal cfDNA copy-number profiles (log₂ ratio) inferred by ichorCNA for a representative patient (M4-05) at diagnosis, 32 days before biochemical progression, and at clinical relapse. Each dot represents a genomic bin, with color indicating the inferred copy-number state (blue = neutral, red = gain, green = loss). Vertical dashed lines mark chromosome boundaries. The annotation above each plot shows the estimated tumor fraction, ploidy, and subclonal fraction. Orange arrows highlight focal copy-number alterations that emerged between diagnosis and relapse, indicating expansion of resistant subclones detectable in cfDNA prior to clinical progression. **(B)** Additional four representative patients demonstrating emergence or enrichment of subclonal copy-number events between diagnosis (Dx) and progression (Prog) in cfDNA. Orange arrows mark new or expanding aberrations absent at diagnosis. Together, these longitudinal cfDNA profiles reveal dynamic subclonal evolution and acquisition of treatment-resistant genomic alterations preceding relapse.

CNA-based tumor-fraction estimates from ichorCNA corroborated these findings. Across all 10 patients, tumor fraction generally increased approaching progression, consistent with rising disease burden. Median tumor fraction rose from 0.0% at nadir to 7.4% at relapse (range 2.6– 40.3%), with nadirs typically occurring several months before clinical progression (median 274 days, Extended Data Fig. 3C). Patients with emergent CNAs exhibited more pronounced increases in tumor fraction than those without (median rise 11.6 vs. 3.6 percentage points), consistent with both greater tumor shedding and the dependence of CNA detection in blood cfDNA on tumor fraction.

## Discussion

Our work shows that moderate-depth (30-40X) cfWGS provides a minimally invasive readout of residual disease dynamics in MM during maintenance therapy, thereby addressing limitations inherent in traditional BM-based approaches. By leveraging personalized baseline mutation profiles from BM WGS, cfWGS demonstrated strong (75-90%) concordance with established MRD assays (MFC and clonoSEQ), and stratified relapse risk at one-year maintenance (HR = 24.14). When diagnostic BM was insufficient or unavailable, a cfDNA-only strategy seeded from baseline blood achieved moderate performance (mAUC 0.79; sensitivity 70%; specificity 90% with fragmentomic features), expanding surveillance eligibility while acknowledging reduced sensitivity near the detection limit.

Although BM MRD assays remain more analytically sensitive at very low disease levels, cfWGS is minimally invasive and easily repeatable, enabling high-frequency, systemic surveillance that mitigates sampling error from patchy or hemodiluted aspirates. In this cohort, cfWGS MRD status at one-year maintenance separated PFS more strongly than marrow MRD (cfWGS HR = 24.14, 95% CI 2.75–211.73; MFC HR = 4.28, 95% CI 0.81–22.1), though event numbers were limited and confidence intervals were wide. This difference likely reflects reduced sampling bias and the broader systemic genomic coverage of cfDNA, which can capture emergent lesions missed by a single-site aspirate. Consistent with this, cfWGS identified marrow-negative but cfDNA-positive cases that later relapsed (e.g., M4-05, M4-33), supporting its ability to detect systemic residual disease missed by localized sampling. Lastly, cfWGS detected subclonal evolution at progression in 50% of profiled patients and signaled rising MRD probabilities a median of 12–19 months before IMWG clinical progression, providing actionable lead time. Together, these properties position cfWGS as a complement to marrow MRD, being less invasive, more repeatable, and providing a more systemic and genomically comprehensive view of disease, supporting its clinical utility for longitudinal MRD surveillance.

Our work builds upon concepts of tumor-informed mutational sampling and fragmentomics by cfWGS (ie MRDetect^30^, MRD-EDGE^37^) by retaining its z-score framework while adding complementary quantitative and biological signals to improve sensitivity and interpretability. Our approach builds on this paradigm but differs in design. We retained a z-score– based framework and introduce interpretable metrics, including the site-hit fraction, mutant-read fraction, and fragmentomic z-scores relative to healthy controls rather than using abstract learned features. We then calibrated a probabilistic classifier for clinical MRD (clonoSEQ/MFC) in MM and implement a plasma-only, baseline-seeded mode for patients who lack marrow WGS. Optimized for moderate-depth (30–40×) sequencing in a hematologic context, our method complements MRD-EDGE by emphasizing interpretability, clinical calibration, and feasibility for serial monitoring in MM.

Response depths achieved with contemporary regimens make sustained MRD the clinically relevant target. In the phase 3 PERSEUS trial, daratumumab plus bortezomib, lenalidomide, and dexamethasone (D-VRd) yielded complete responses in nearly 90% of transplant-eligible patients and BM MRD negativity in roughly three quarters, with four-year PFS of 84% versus 68% for VRd alone.^38,39^ Similar findings from GRIFFIN and CASSIOPEIA showed concordant patterns with daratumumab-containing quadruplets.^3,40^ Trials have operationalized MRD durability as a treatment decision point. In MASTER, patients with standard-risk disease discontinued therapy after two consecutive MRD-negative results, while high-risk patients frequently relapsed after MRD loss;^41^ in PERSEUS, daratumumab maintenance was stopped after two years of sustained BM MRD negativity.^38,39^ These designs make a blood-based assay capable of serially detecting MRD loss or durability immediately actionable.

Recent prospective studies using blood-based MRD detection using next-generation flow (NGF) or mass spectrometry (MS) add independent prognostic value and complement marrow MRD during maintenance.^42,43^ Within this landscape, cfWGS contributes orthogonal information including genome-wide mutation and copy-number signals, the ability to detect structural variation (with depth-dependent sensitivity), and fragmentomic features, providing molecular context that protein- or cell-based assays cannot. As bispecific antibodies enter routine practice, antigen escape from targeted receptors (BCMA, GPRC5D) is an increasingly recognized resistance mechanism that can shape sequencing of TCRT. Genomic studies at relapse have documented biallelic *TNFRSF17* (BCMA) loss and *GPRC5D* modulation after targeted immunotherapies.^44,45^ cfWGS can potentially detect such target-locus alterations in plasma, flagging antigen-negative relapse and informing target switching during serial monitoring. These findings highlight the clinical potential of blood-based MRD tools for integrating cfWGS into MRD-guided trial designs.

We also evaluated cfDNA fragmentation metrics (median fragment length, short-to-long fragment ratios, coverage biases at regulatory regions), which correlate with tumor burden in multiple cancers.^31,46–48^ In our cohort, selected fragmentomic features correlated significantly with disease burden indicators (clonoSEQ, residual plasma-cell percentages, tumor fraction). However, classification performance based solely on these fragmentomic features was modest (best AUC=0.63), contrasting with higher AUCs (0.78–0.94) in solid-tumor studies employing extensive feature sets and machine-learning approaches.^31,46,47^ Our limited feature set and smaller cohort size restricted predictive power. Notably, when integrated with mutation-based features, fragmentomics improved blood-based classification performance, underscoring its value as a complementary signal. Thus, broader feature sets, deeper sequencing, and larger cohorts, similar to prior studies,^31,46,47^ are necessary to enhance the standalone accuracy of fragmentomics-based MRD classification in MM. Future studies should evaluate whether integrating expanded fragmentomic feature sets meaningfully improves cfDNA-based MRD detection.

Our study has several limitations. The modest cohort size limits statistical power for detailed subgroup analyses and survival correlations. Additionally, focal or low-shedding residual disease confined to the marrow may escape cfDNA detection, risking false negatives. Infrequent sampling intervals also limited temporal resolution for tracking subclonal evolution. More frequent sampling (e.g., quarterly) could refine the timing of molecular relapse and enable earlier intervention, while deeper sequencing may reveal emergent somatic mutations with greater sensitivity. Larger multicenter studies with standardized sampling and long follow-up are needed to confirm prognostic value and define optimal cfWGS thresholds for clinical decision-making. Technical refinements, such as deeper sequencing or improved error profiling,^49^ broader fragmentomic feature sets coupled to machine-learning classifiers,^47^ and integration of broader orthogonal signals (such as cfDNA methylation or immune-profiling)^50^, should lower the detection floor and clarify optimal assay combinations. Beyond MM, cfWGS’s simultaneous capture of mutations, CNAs, and SVs positions it ideally for other cancers with significant ctDNA shedding, limited biopsy feasibility, or tumor heterogeneity, including diffuse large B-cell lymphoma^51^ and high-grade serous ovarian carcinoma.^52^

In conclusion, our findings establish moderate depth cfWGS as a sensitive, clinically informative approach for MRD detection in MM. With an empirical limit of detection of 0.011% VAF, high concordance with established assays, and the ability to detect subclonal evolution, cfWGS addresses critical gaps in MRD monitoring. Prospective validation in larger cohorts will clarify its clinical impact and inform optimal integration into patient management strategies. By enabling minimally invasive longitudinal monitoring, early relapse detection, and improved understanding of tumor biology, cfWGS offers a capable framework to support MRD-adapted care focused on durability of remission and improving patient outcomes in MM and other malignancies.

## Data Availability

Code is available on GitHub. The repository includes scripts to reproduce model training/evaluation and figure panels. Human genomic data underlying this study is currently being deposited in a controlled-access repository (EGA). Summary-level data and per-figure source data are provided as Supplementary Tables and in the Supplementary Information. Any additional materials supporting the findings are available from the corresponding authors upon request.

https://github.com/pughlab/cfWGS-MM-MRD

## Acknowledgements

We gratefully acknowledge the Terry Fox Research Institute (TFRI) M4 Study and the Paula and Rodger Riney Foundation for their generous support which made this project possible. This work was also funded through the Ontario Institute for Cancer Research Genomics Program with support from the Government of Ontario and benefited from infrastructure provided by the Canada Foundation for Innovation (CFI) through the Leaders Opportunity Fund (CFI #32383 and #38401), the Ontario Ministry of Research and Innovation, and the Ontario Research Fund Small Infrastructure Program. We also thank the Marathon of Hope Cancer Centres Network (MOHCCN) for supporting the tumor sequencing of the IMMAGINE cases. We are also grateful to the SPORE team at Mayo Clinic Rochester and Mayo Clinic Arizona for collaborative input, and to members of the Pugh and Trudel laboratories for their ongoing discussions and support. T.J.P. is the Canada Research Chair in Translational Genomics and receives funding from the Gattuso-Slaight Personalized Cancer Medicine Fund as well as an OICR Senior Investigator Award. We gratefully acknowledge the contributions of the Princess Margaret Genomics Centre staff and the UHN High-Performance Computing and Bioinformatics Core.

## Author Contributions

D.D.A. performed computational and statistical analyses, prepared figures, drafted and revised the manuscript, directed sample selection and submission, and designed and implemented the machine-learning framework, and assisted with the study design with input from T.J.P. and S.T. T.J.P. and S.T. conceived the study, supervised the research, secured funding, and provided overall leadership. J.E. and S.P. processed cfDNA biospecimens, including DNA extractions. A.W. and S.S. processed bone marrow biospecimens and coordinated clinical MRD testing with A.R. E.N.W. and S.A. assisted with IMMAGINE sample processing. D.S.S. curated clinical and sample data. J.P.B., E.B., A.K.S., C.B.d.C., and S.C. contributed to data interpretation. D.W., I.S., K.S., S.K., and A.R. enrolled patients and provided biospecimens and clinical data. All authors reviewed and approved the final manuscript.

### Competing Interests

D.W. has received honoraria from Janssen, Novartis, Forus Therapeutics, Sanofi, Antengene, Pfizer, and GlaxoSmithKline. K.S. has received honoraria from Novartis, Janssen, GlaxoSmithKline, Bristol Myers Squibb, Gilead, and Sanofi. S.T. reports honoraria from Janssen, Pfizer, Kite, GlaxoSmithKline, Sanofi, Roche, and K36 Therapeutics; consultancy for GlaxoSmithKline and Roche; and research funding from Janssen, Bristol Myers Squibb, Pfizer, Roche, and K36 Therapeutics. S.K. reports consulting with no personal payment for AbbVie, Amgen, ArcellX, Beigene, Bristol Myers Squibb, Carsgen, GSK, Janssen, K36, Moderna, Pfizer, Regeneron, Roche-Genentech, Sanofi, and Takeda; personal consulting payments from CVS Caremark and BD Biosciences; and clinical trial support to their institution from AbbVie, Amgen, AstraZeneca, Bristol Myers Squibb, Carsgen, GSK, Gracell Bio, Janssen, Oricell, Roche-Genentech, Sanofi, Takeda, and Telogenomics. T.J.P. reports consultancy for Roche, AstraZeneca, Merck, and Chrysalis Biomedical Advisors; research funding from Roche, Genentech, and AstraZeneca; and patents/royalties with the University Health Network and Dynacare. S.P. also reports royalties with the University Health Network and Dynacare. All disclosures are outside the scope of the submitted work. The remaining authors declare no competing interests.

## Extended Data Figures

**Extended Data Figure 1:**
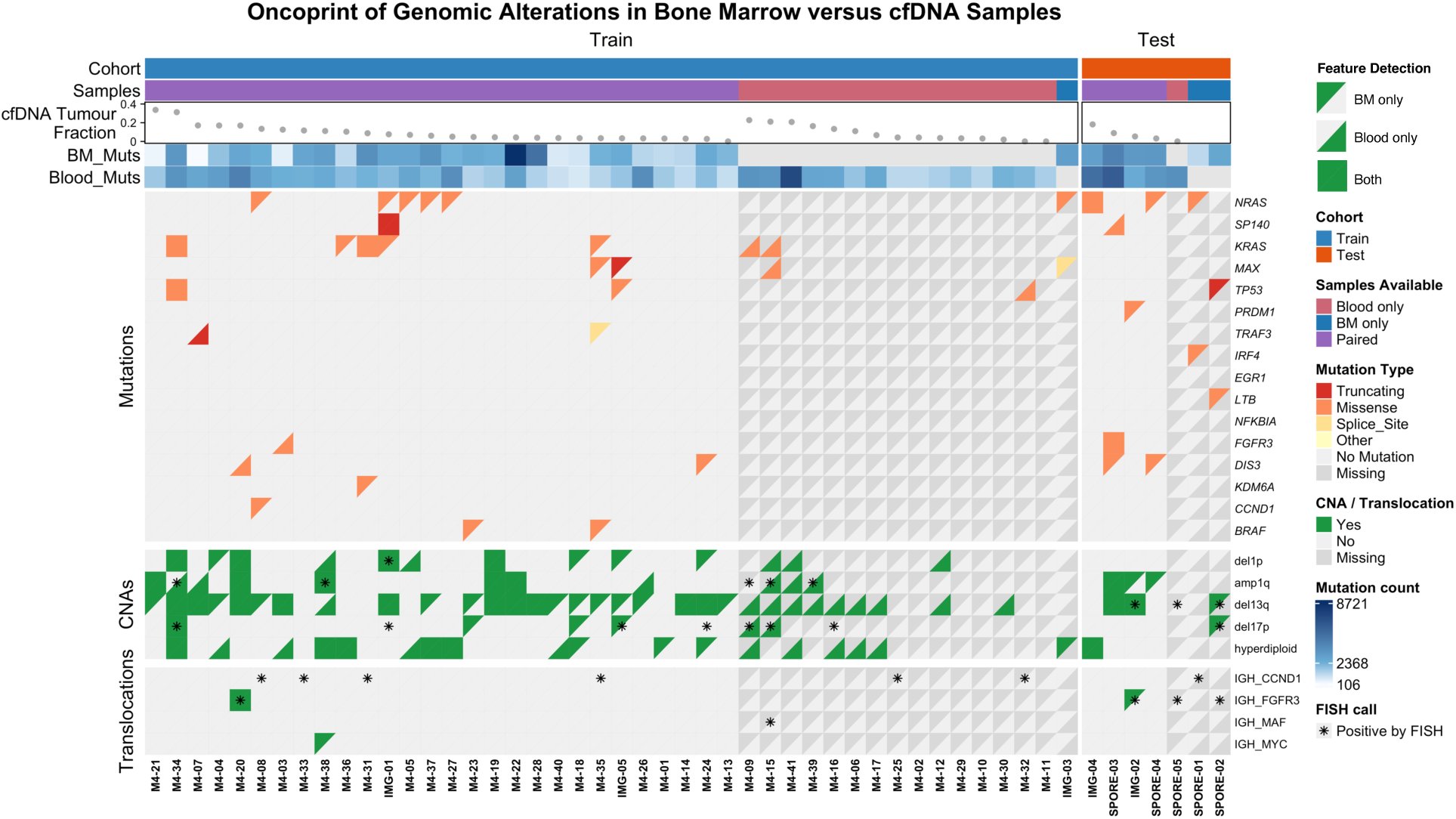
Baseline genomic landscape of bone marrow and cfDNA samples. Genomic characterization at baseline for the patient cohorts. Upper bars indicate cohort assignment and sample type availability (bone marrow, cfDNA, or paired). The scatter plot depicts estimated CNA-based cfDNA tumor fractions (from ichorCNA^53^). Heatmap panels detail the mutational status of recurrently altered myeloma genes (top), and key copy-number alterations (CNAs), including deletions (del1p, del13q, del17p), amplifications (amp1q), and hyperdiploidy (middle). The lower panel shows IGH translocation status, where confirmed events by fluorescence in situ hybridization (FISH) are indicated by asterisks (*).

**Extended Data Figure 2:**
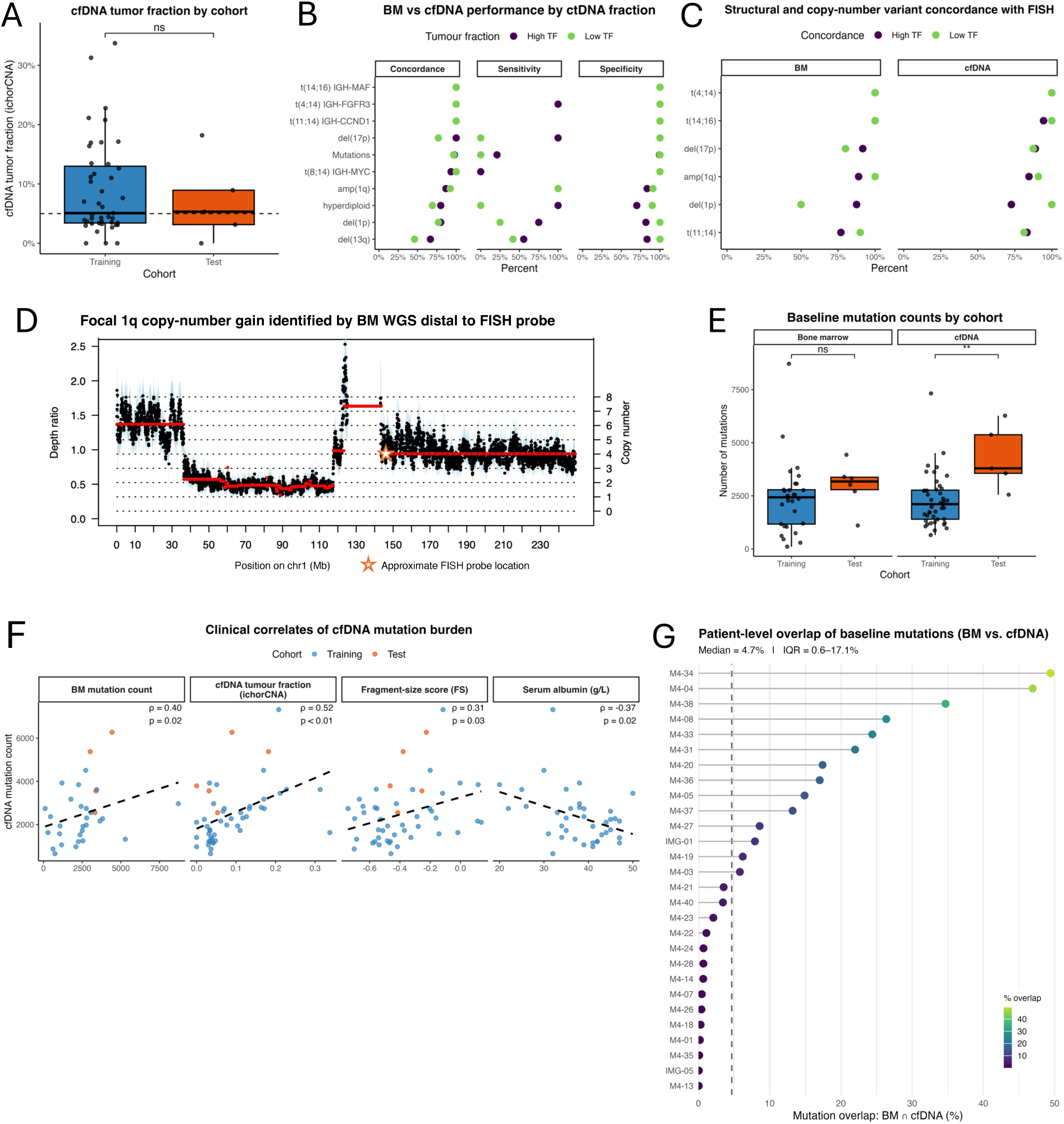
Concordance between bone marrow and cfDNA genomic profiles at baseline. **(A)** Estimated cfDNA tumor fraction (ichorCNA) at baseline, stratified by training and test cohorts. **(B)** Performance of cfDNA in detecting bone marrow (BM) genomic features (translocations and CNAs), stratified by cfDNA tumor fraction. Concordance, sensitivity, and specificity are shown across increasing tumor fraction thresholds. **(C)** Comparison of structural variant and CNA detection by sequencing versus FISH, for both BM and cfDNA. Overall concordance is shown, stratified by cfDNA tumor fraction. **(D)** Bone marrow WGS copy-number profile generated with Sequenza showing a focal 1q gain located distal to the 1q21 FISH probe. **(E)** Baseline mutation counts in BM and cfDNA samples by cohort. **(F)** Correlations between cfDNA mutation burden and clinical/molecular features, including BM mutation count, cfDNA tumor fraction, cfDNA fragment-size score (FS), and serum albumin. Dashed lines represent fitted regression models. **(G)** Patient-level concordance of mutation calls between BM and cfDNA at baseline. The percentage of overlapping mutations is shown per patient, colored by overlap proportion. These analyses demonstrate the degree of molecular concordance between BM and cfDNA across cohorts and highlight factors that influence the sensitivity of cfDNA for capturing baseline genomic alterations in multiple myeloma.

**Extended Data Figure 3:**
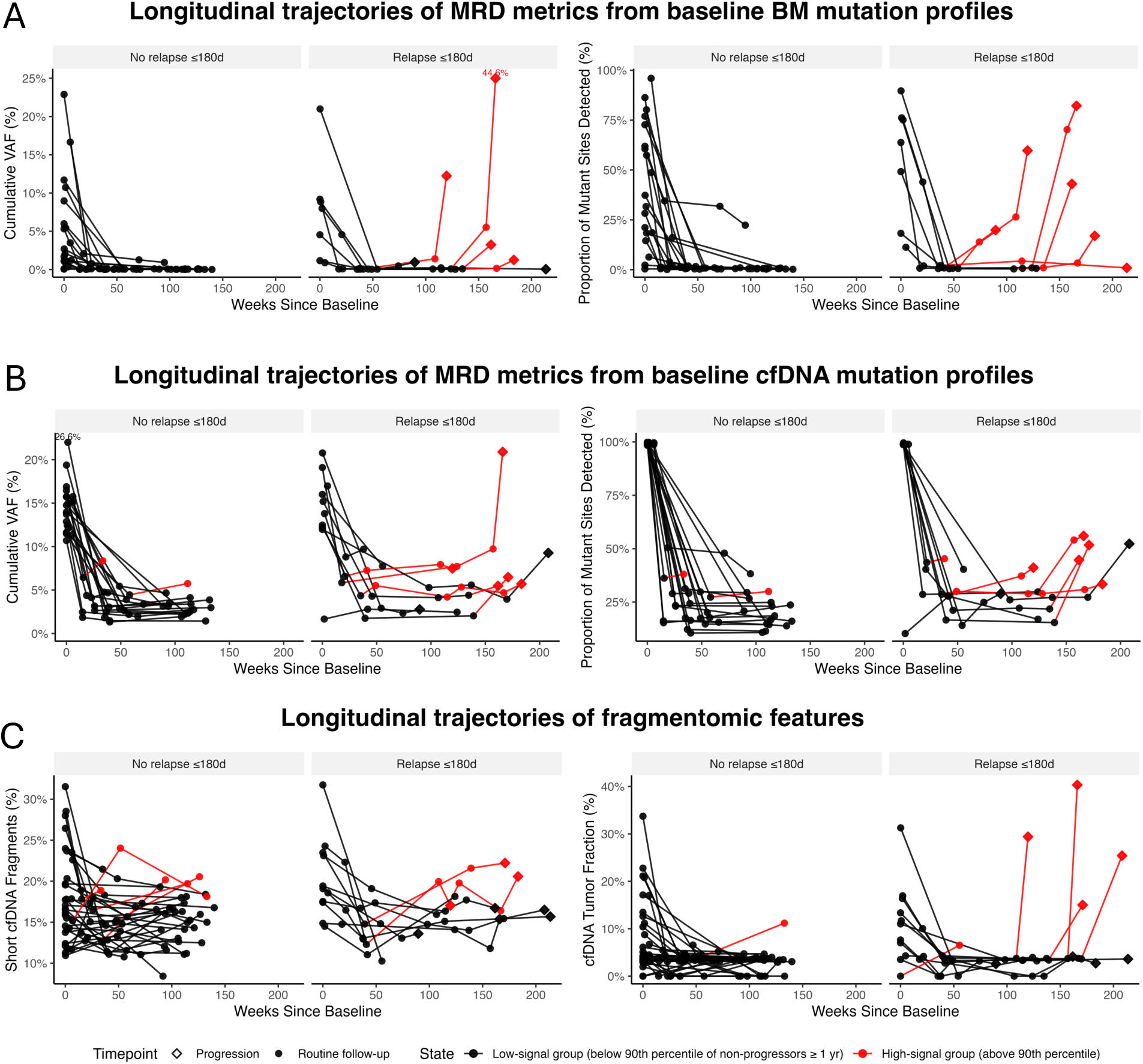
Additional longitudinal MRD dynamics from tumor- and blood-informed mutation profiles and fragmentomic features. **(A)** Longitudinal trajectories of MRD metrics derived from baseline bone marrow (BM) mutation profiles, including cumulative variant allele frequency (VAF) and the proportion of mutant sites detected from baseline. **(B)** Corresponding trajectories for metrics derived from baseline peripheral blood (PB) cfDNA mutation profiles. **(C)** Longitudinal dynamics of fragmentomic features in cfDNA, including short-fragment proportion and cfDNA tumor fraction (estimated by ichorCNA). Each line represents an individual patient. Patients are stratified by relapse within 180 days of the last available sample versus no relapse. Points are colored red when the sample value exceeded the 90th percentile of non-progressors ≥1 year from baseline. These additional features complement the primary metrics in Figure 2 and reinforce the prognostic utility of cfDNA-based mutation and fragmentation signatures for sensitive, non-invasive MRD monitoring in multiple myeloma.

**Extended Data Figure 4:**
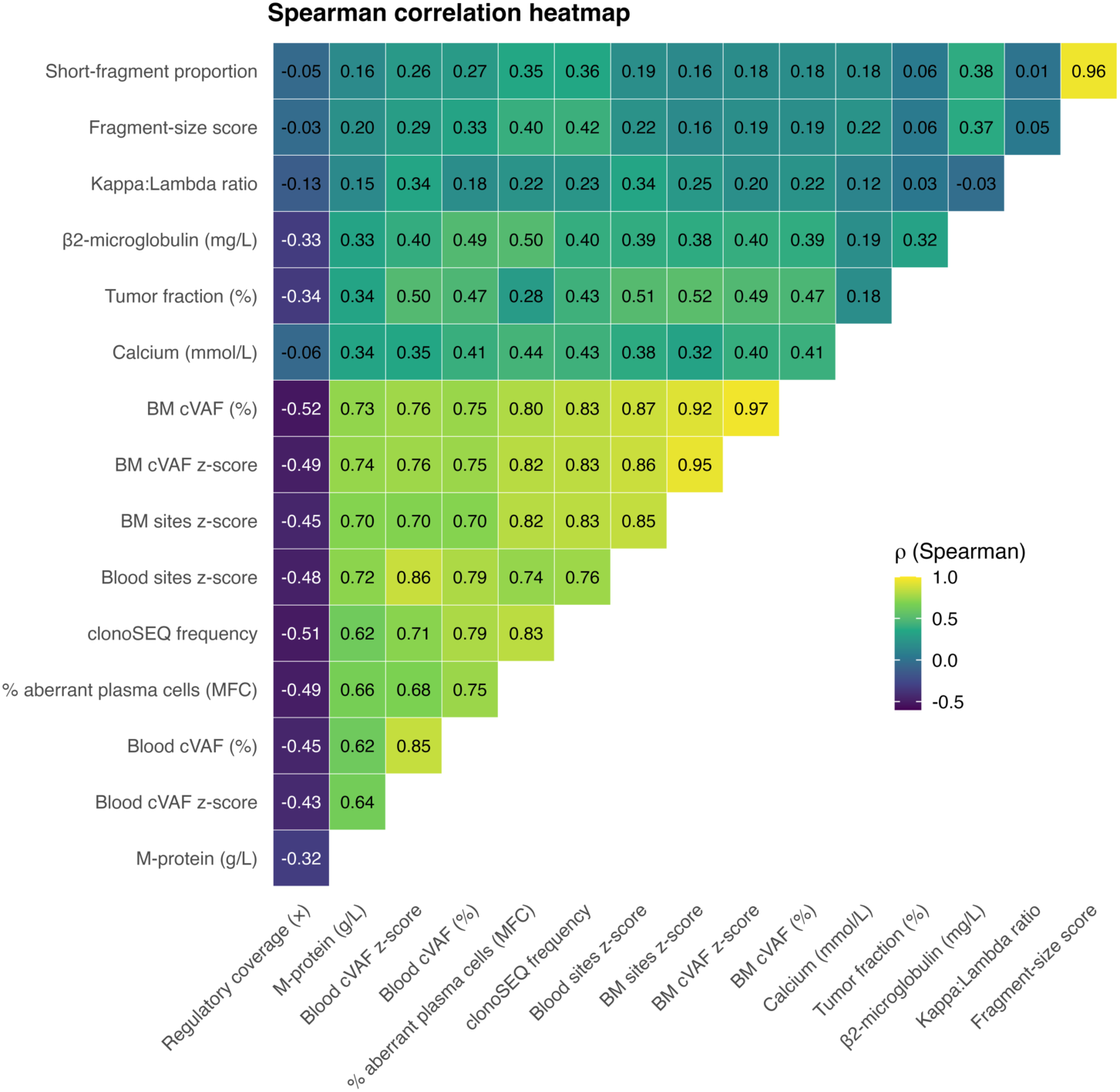
Correlation of cfDNA- and BM-derived genomic features with clinical and laboratory parameters. Spearman correlation heatmap comparing cfDNA-based, bone marrow-based, and clinical MRD metrics with disease-related laboratory features and tumor burden indicators. Features include short-fragment proportion, cfDNA fragment-size score, κ:λ ratio, β2-microglobulin, tumor fraction, serum calcium, clonoSEQ frequency, M-protein, and percentage of aberrant plasma cells by MFC. cfDNA- and BM-derived features (cVAF, Z-scores, and number of sites detected) are positively correlated with established myeloma disease burden markers such as β2-microglobulin, clonoSEQ MRD, and M-protein concentration. Notably, clonoSEQ frequency and MFC were among the strongest correlates of both cfDNA and BM signal intensity. Color intensity reflects correlation strength, with yellow indicating a strong positive correlation and purple indicating inverse correlation. This analysis illustrates how cfDNA-based genomic and fragmentomic measures align with known markers of disease activity in multiple myeloma.

**Extended Data Figure 5:**
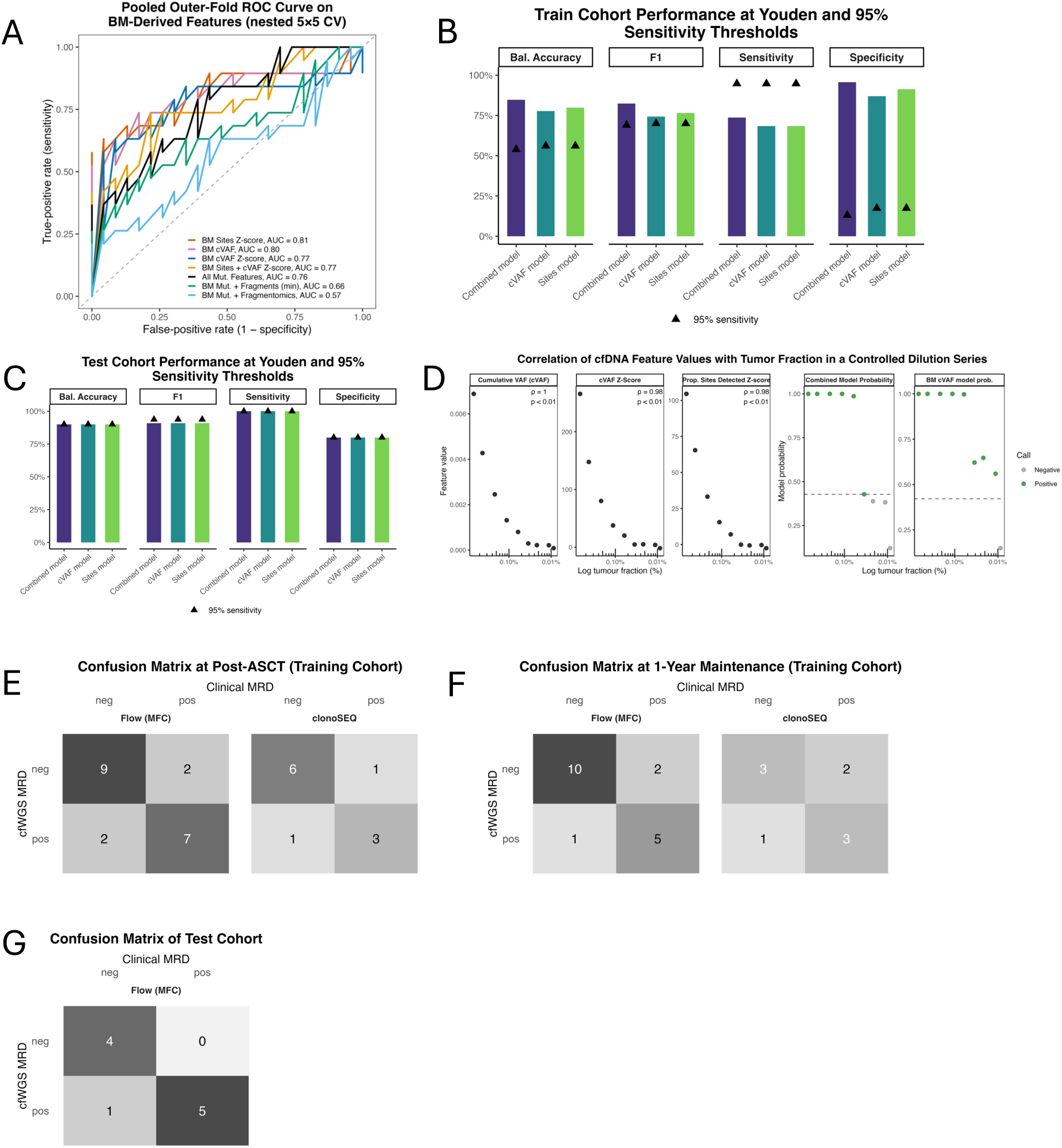
Performance and validation of cfDNA-based MRD classifiers using bone marrow-derived mutation lists. **(A)** Cross-validated receiver operating characteristic (ROC) curves demonstrating predictive accuracy of various cfDNA-based classifiers using BM mutational and fragmentomic features. Area under the ROC curves (AUCs) represents discriminative performance based on nested 5×5-fold cross-validation. Legend mapping: BM Sites Z-score = z-score of the proportion of baseline loci with ≥1 mutant read; BM cVAF = pooled mutant read fraction across baseline loci; BM cVAF Z-score = standardized cVAF; All Mut Features = Sites Z-score + cVAF + cVAF Z-score; BM + Fragmentomics = All Mut Features + fragment score (FS), mean GC-corrected coverage at MM regulatory sites, short-fragment proportion, and cfDNA tumor fraction (ichorCNA); BM + Fragments (min) = All Mut Features + FS + mean coverage only. **(B)** Training-cohort performance at Youden (bars) versus fixed 95% sensitivity (triangles) thresholds. Balanced accuracy, F₁ score, sensitivity, and specificity are shown for three models: Combined (purple), cVAF z-score to healthy controls (teal), and the proportion of sites detected z-score relative to healthy controls (green). **(C)** Test-cohort performance at Youden (bars) and 95% sensitivity (triangles). Same metrics and models as in (A), evaluated on the held-out test set. **(D)** Correlation of cfDNA features and MRD model probabilities with tumor fraction in a controlled dilution series. Feature values and model-derived MRD probabilities from cfWGS were evaluated across serial dilutions of tumor DNA to benchmark sensitivity at low tumor fractions. Bone marrow–derived metrics (cumulative VAF, cVAF z-score, and proportion of mutant sites detected) and corresponding model probabilities show strong correlation with tumor fraction, confirming the reliability of cfDNA-based MRD detection at ultra-low tumor burden. **(E)** Confusion matrices at the post-ASCT time point in the training cohort. Left: comparison of cfWGS MRD calls to flow cytometry (MFC). Right: cfWGS versus clonoSEQ. Cells show TN, FP, FN, and TP counts. **(F)** Confusion matrices at the 1-year maintenance time point (training cohort). Left: cfWGS versus MFC. Right: cfWGS versus clonoSEQ. **(G)** Confusion matrix in the independent test cohort. cfWGS MRD calls versus flow cytometry (MFC) at the matched time point. This figure highlights the concordance between cfWGS-based MRD classifiers and standard clinical MRD assays (flow cytometry and clonoSEQ), demonstrating strong performance across folds and cohorts. These results support the use of cfDNA-derived metrics as a reliable, minimally invasive approach for detecting residual disease at clinically relevant time points.

**Extended Data Figure 6:**
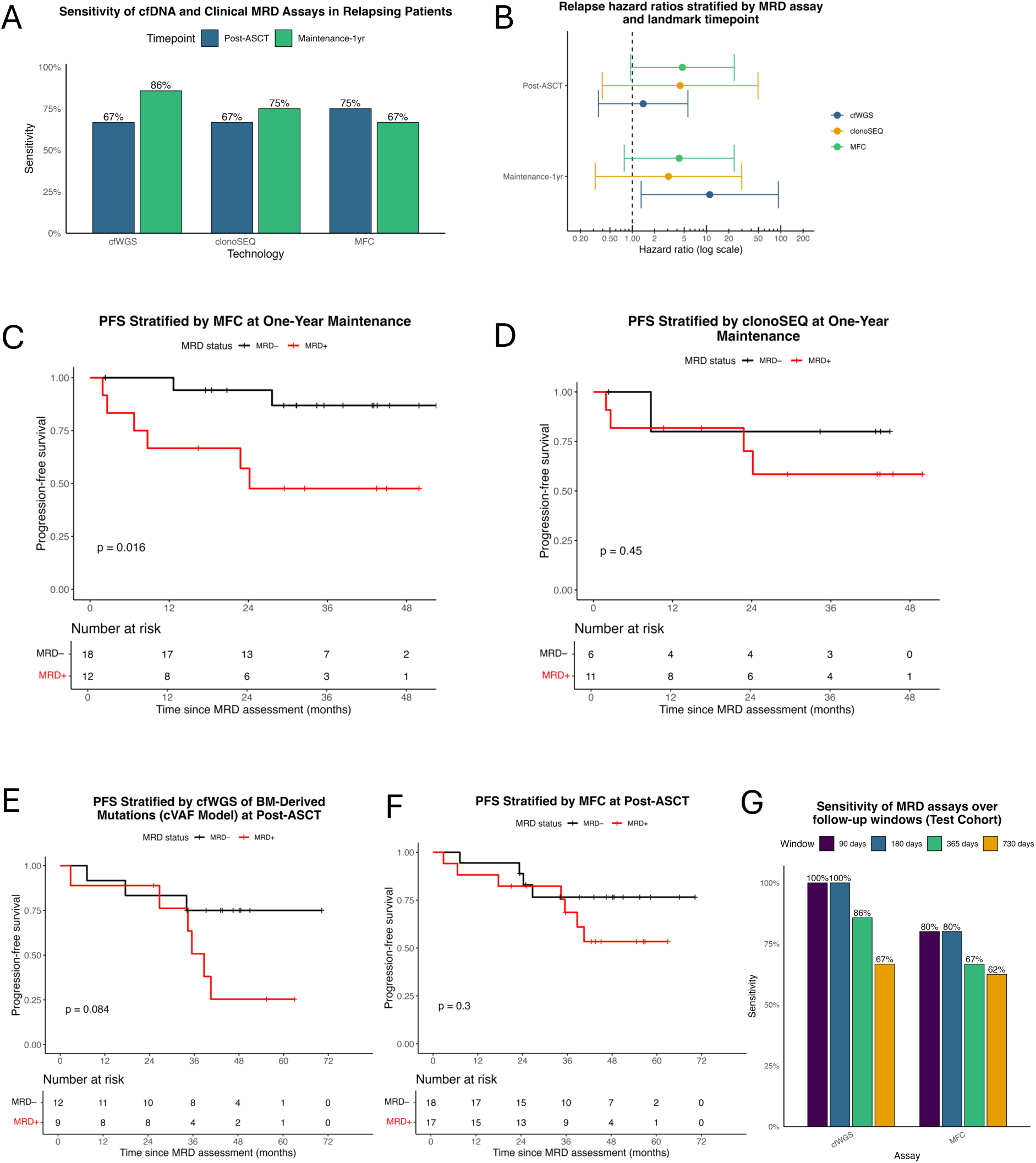
Sensitivity and prognostic value of MRD assays for detecting relapse. **(A)** Sensitivity of cfWGS, clonoSEQ, and MFC for detecting MRD positivity among patients who eventually relapsed, stratified by time point (post-autologous stem-cell transplant [ASCT] and 1-year maintenance). **(B)** Hazard ratios for clinical relapse, stratified by MRD status (positive vs. negative), MRD assay (cfWGS, clonoSEQ, MFC), and landmark time point. Horizontal bars represent 95% confidence intervals; the vertical dashed line marks HR = 1. **(C, D)** Kaplan–Meier curves showing time to progression stratified by MRD status as determined by MFC (C) and clonoSEQ (D) in a landmark analysis at one year of maintenance therapy. Numbers at risk are shown below each time point. **(E, F)** Kaplan–Meier curves showing time to progression from the post-ASCT time point, stratified by MRD status as determined by cfWGS using the cVAF model from BM-derived mutation lists (E) and by MFC (F). Numbers at risk are shown below each time point. **(G)** Sensitivity of each MRD assay (cfWGS, clonoSEQ, MFC) across multiple follow-up windows (90–720 days) in the test cohort. While panels A–F examine landmark-based associations with progression, this panel assesses dynamic detection sensitivity over rolling follow-up intervals for patients without consistent sampling, highlighting cfWGS performance across clinically relevant timescales. These results highlight the varying sensitivity and prognostic performance of each MRD assay at clinically relevant time points, with cfWGS and MFC providing complementary insights into relapse risk.

**Extended Data Figure 7:**
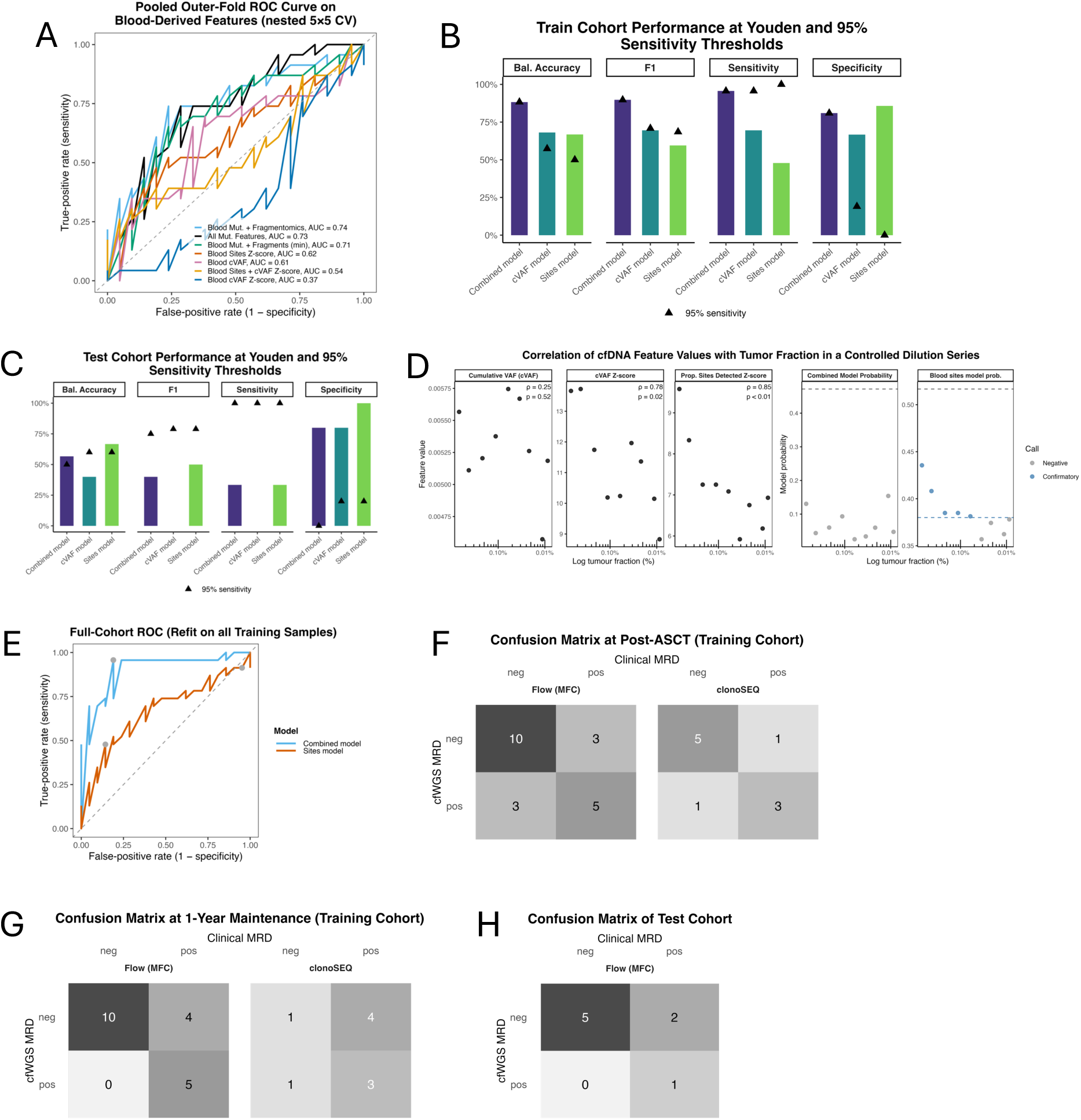
Performance and validation of cfDNA-based MRD classifiers using blood cfDNA-derived mutation lists. **(A)** Cross-validated receiver operating characteristic (ROC) curves demonstrating predictive accuracy of various cfDNA-based classifiers using BM mutational and fragmentomic features. Area under the ROC curves (AUCs) represents discriminative performance based on nested 5×5-fold cross-validation. Legend mapping: Blood Sites Z-score: z-scored proportion of baseline loci with ≥1 mutant read; Blood cVAF: pooled mutant read fraction across baseline loci; Blood cVAF Z-score: standardized cVAF; All Mut Features: Sites Z-score + cVAF + cVAF Z-score; Blood + Fragmentomics: All Mut Features + fragment score (FS) + mean GC-corrected coverage at MM regulatory sites + short-fragment proportion + cfDNA tumor fraction (ichorCNA); Blood + Fragments (min): All Mut Features + FS + mean coverage only. **(B)** Training-cohort performance at Youden (bars) versus fixed 95% sensitivity (triangles) thresholds. Balanced accuracy, F₁ score, sensitivity, and specificity are shown for three models: Combined (purple), cVAF z-score to healthy controls (teal), and the proportion of sites detected z-score relative to healthy controls (green). **(C)** Test-cohort performance at Youden (bars) and 95% sensitivity (triangles). Same metrics and models as in (A), evaluated on the held-out test set. **(D)** Correlation of cfDNA features and MRD model probabilities with tumor fraction in a controlled dilution series. Feature values and model-derived MRD probabilities from cfWGS were evaluated across serial dilutions of tumor DNA to benchmark sensitivity at low tumor fractions. cfDNA–derived metrics (cumulative VAF, cVAF z-score, and proportion of mutant sites detected) and corresponding model probabilities show strong correlation with tumor fraction. **(E)** Pooled ROC curves from nested 5×5-fold cross-validation for the Combined Model using cfDNA-derived mutation and fragmentomic features (blue) and Sites Model based on the z-score of the proportion of mutant sites detected relative to healthy controls (orange). Circles mark Youden thresholds and the high sensitivity screen threshold for the Sites Model. **(F)** Confusion matrices at the post-ASCT landmark in the training cohort: cfWGS MRD calls versus flow cytometry (MFC, left) and versus clonoSEQ (right). Cells display true negatives, false positives, false negatives, and true positives. **(G)** Confusion matrices at the 1-year maintenance landmark in the training cohort, comparing cfWGS MRD to MFC (left) and to clonoSEQ (right). **(H)** Confusion matrix in the independent test cohort comparing cfWGS MRD calls to MFC. These panels demonstrate that classifiers built from blood cfDNA mutation lists achieve moderate cross-validated performance, high concordance with gold-standard MRD assays, and reproducible accuracy in both training and test cohorts.

**Extended Data Figure 8:**
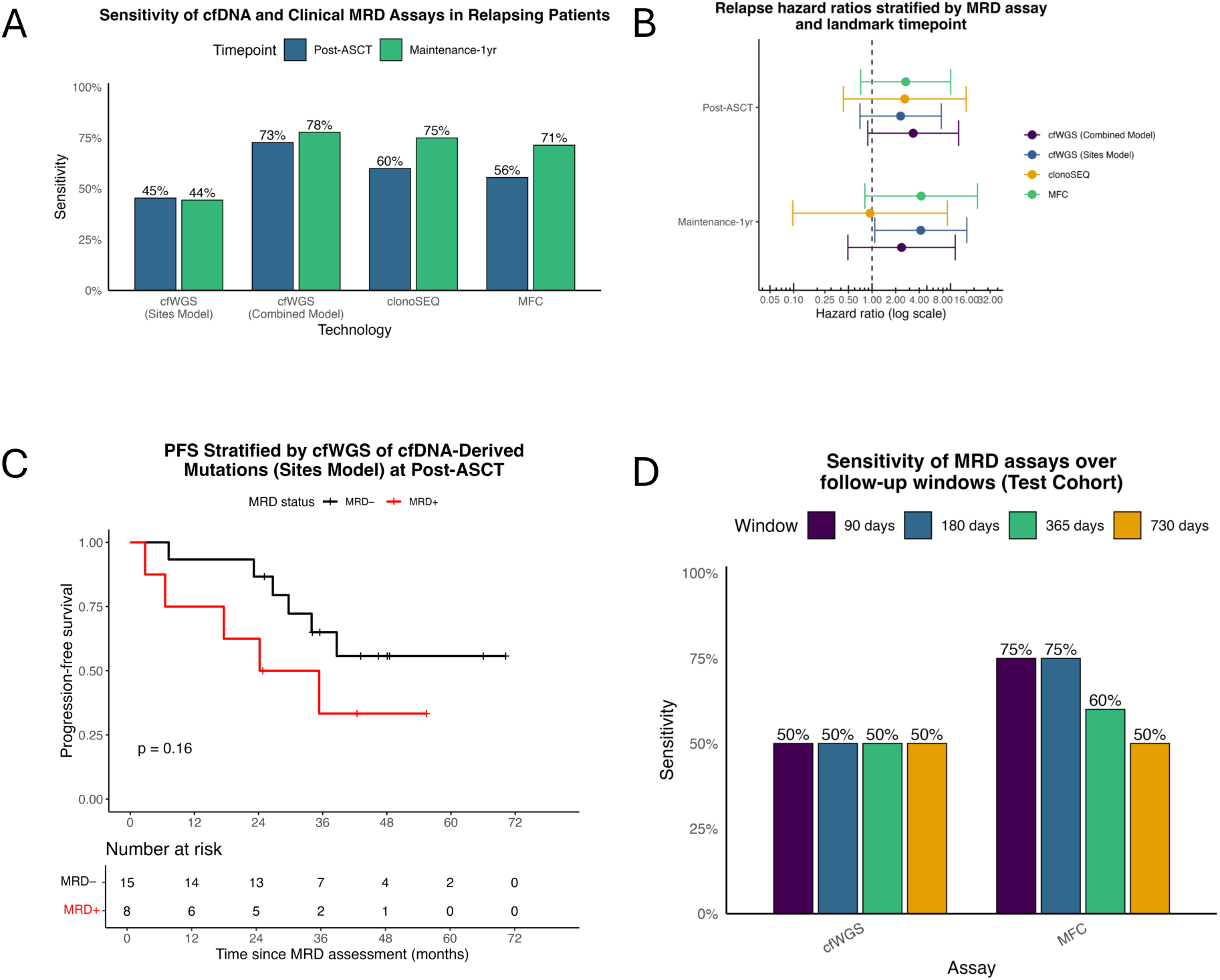
Sensitivity and prognostic value of blood cfDNA-based cfWGS for relapse detection. **(A)** Sensitivity of MRD detection by PB-derived cfWGS, clonoSEQ, and MFC among patients who later relapsed, stratified by post-ASCT and 1-year maintenance time points. **(B)** Relapse hazard ratios for MRD-positive versus MRD-negative calls, by assay and landmark time point. Error bars denote 95 % confidence intervals; the vertical line marks HR = 1. **(C)** Kaplan–Meier curves for progression-free survival stratified by the Sites Model (based on the z-score of the proportion of mutant sites known from baseline cfDNA detected relative to healthy controls) MRD status at the post-ASCT landmark. Numbers at risk are shown below each time point. **(D)** Sensitivity of each MRD assay (PB-derived cfWGS and MFC) over rolling follow-up windows (90, 180, 365, 730 days) in the test cohort using the Sites Model. This figure highlights that PB-derived cfWGS MRD detection is sensitive at key treatment milestones, stratifies progression-free survival, and maintains predictive performance across extended follow-up, supporting its value as a noninvasive tool for relapse monitoring.

**Extended Data Figure 9:**
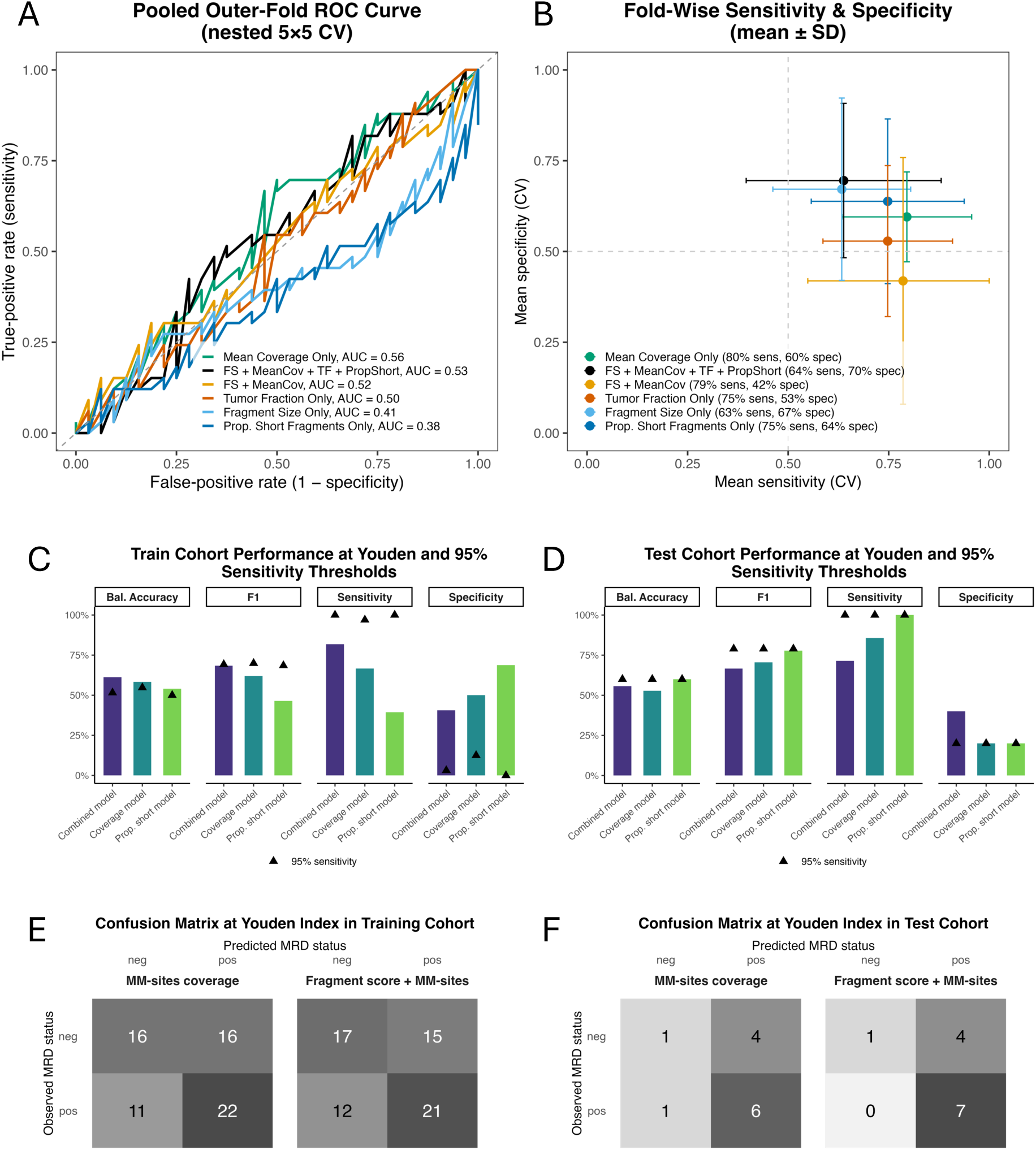
Performance of fragmentomics-only models for MRD classification without a baseline sample. **(A)** Pooled ROC curves from nested 5×5-fold cross-validation for six fragmentomic-only models: mean coverage at myeloma regulatory sites (MeanCov), fragment-size score (FS), tumor fraction (TF), proportion of short fragments (PropShort), and two multi-feature combinations (Fragmentomics_min is the fragment score and mean coverage combined, and fragmentomics_full is all fragment features combined). **(B)** Fold-wise mean sensitivity versus specificity (±1 SD across outer CV folds) for each model; the mean coverage at myeloma regulatory sites only model attained the best trade-off (mean sensitivity 80%, specificity 60%; pooled AUC = 0.56). **(C, D)** Training (C) and independent test (D) cohort performance for the top three selected models (Combined FS + MeanCov (purple), MeanCov only (teal), and PropShort only (green)) showing balanced accuracy, F₁ score, sensitivity, and specificity at the Youden index (bars) and fixed 95% sensitivity (triangles). **(E, F)** Confusion matrices at the Youden threshold for MeanCov-only (left) and Combined FS + MeanCov (right) models in the training (E) and test (F) cohorts, with true/false positive and negative counts. This figure shows that although combining fragment-size score with site-specific coverage modestly improves discrimination, overall fragmentomic features alone yield only moderate sensitivity, specificity, and AUC, underscoring the need to integrate them with mutation-based or other orthogonal MRD metrics.

## Supplementary Table Legends

**Supplementary Table 1: Patient treatment histories and sample collection timing**

Treatment details for all patients included in the study, including induction regimen, transplant status, maintenance therapy, and timing of sample collection relative to treatment milestones.

**Supplementary Table 2: Concordance of WGS calls between sample types and with clinical FISH results**

(A) Pairwise concordance and performance between bone marrow and cfDNA performance by genomic feature and tumor fraction (TF) group (High TF, Low TF, All), using BM WGS as the reference. For each feature (aggregated categories: All Translocations, All CNAs, All Mutations, and individual events), the table reports: n = number of samples; TP = true positives; TN = true negatives; FP = false positives; FN = false negatives; and derived metrics including sensitivity, specificity, accuracy, positive predictive value (PPV), negative predictive value (NPV), F1 score, false discovery rate (FDR), false omission rate (FOR), Matthews correlation coefficient (MCC), and Jaccard index.

(B) Agreement of BM WGS and cfDNA WGS with clinical FISH for CNAs and translocations (events: amp1q, del1p, del13q, del17p, hyperdiploidy; IGH–CCND1, IGH–FGFR3, IGH–MAF, IGH–MYC). For each source and event, the table lists TP, TN, FP, FN, sensitivity, specificity, concordance (accuracy), PPV, and NPV. NA denotes metrics not estimable due to zero denominators.

(C) Probe-level comparison of BM WGS and cfDNA WGS with clinical FISH for copy number alterations (events: amp1q, del1p, del17p). For each source and event, the table lists TP, TN, FP, FN, sensitivity, specificity, concordance (accuracy), PPV, and NPV. Copy-number calls were restricted to the cytoband containing the FISH probe; in cfDNA, ichorCNA regions not directly callable due to low mappability were substituted with the nearest available segment. NA denotes metrics not estimable due to zero denominators.

(D) Per-sample comparison of FISH and WGS calls across paired BM and plasma cfDNA. The table lists patient identifier, timepoint, cohort, cfDNA tumor fraction, and BM ploidy estimate, followed by event-level calls for FISH (amp1q, del1p, del17p, t(4;14), t(11;14), t(14;16)) and corresponding WGS results. For each event, both arm-level and probe-level WGS classifications are shown for BM and cfDNA. Boolean columns (“is_altered”) indicate concordance with FISH, taking a value of 1 only when the WGS call matched the FISH-reported alteration (e.g., deletion detected as a loss).

**Supplementary Table 3: Correlations among fragmentomic, genomic, and clinical features**

Correlation matrix of all evaluated DNA and clinical features. Values reflect Spearman correlation coefficients (ρ) with Benjamini-Hochberg (BH) adjusted *p*-values for multiple testing correction. Pairwise correlations were computed using all available sample pairs (*n* = number of non-missing pairs).

**Supplementary Table 4: Outer-fold cross-validation performance of all MRD classifiers**

Summary of classification performance (mean ± SD across outer folds) for each model evaluated in the training cohort. Metrics include mean AUC across folds, SD AUC across folds, pooled AUC with 95% CI, partial AUC at 90% specificity (pAUC90), as well as sensitivity, specificity, positive predictive value (PPV), negative predictive value (NPV), F₁ score, balanced accuracy, at the Youden Index, as well as metrics at 95% specificity, 95% sensitivity, plus Brier score and average training/test set sizes per fold.

**Supplementary Table 5: Training-cohort performance of all models refit on the full dataset**

Performance metrics for all classifiers when refit on the entire training dataset. For each model, metrics are reported at three thresholds: Youden index, 95% specificity, and 95% sensitivity. Reported statistics include threshold value, sensitivity, specificity, PPV, NPV, accuracy, balanced accuracy, precision, and F₁ score.

**Supplementary Table 6: Test-cohort performance of all MRD classifiers**

Same performance metrics as in Supplementary Table 5, evaluated on the test set. Results reflect the generalizability of bone marrow–derived, peripheral blood–derived, and fragmentomics-only classifiers at Youden, 95% specificity, and 95% sensitivity thresholds.

**Supplementary Table 7: Dilution-series cohort performance of all MRD classifiers**

(A) Correlations of different evaluated mutation and fragmentomic features and models against the expected tumor fraction measured from the dilution series

(B) Full table used to generate (A), including model probabilities, calls, and other evaluated mutation and fragmentomic metrics on each sample from the dilution series.

**Supplementary Table 8: Per-sample comparison of cfWGS and clinical MRD calls**

Per-sample comparison of model probabilities, binary cfWGS calls, and matched clinical MRD metrics across technologies and cohorts for the primary bone marrow (BM) and blood cfDNA classifiers. Sheet A: Training cohort, BM-derived mutation lists (cVAF Model). Sheet B: Test cohort, BM-derived mutation lists. Sheet C: Training cohort, blood cfDNA-derived mutation lists (Sites Model). Sheet D: Test cohort, blood cfDNA-derived mutation lists.

**Supplementary Table 9: Time-window performance of relapse prediction models in the test cohort.**

BM and blood cfDNA MRD models and MFC were evaluated for their ability to predict clinical progression within fixed time windows (90, 180, 365, and 730 days). Each row shows the assay tested, number of samples and patients included, confusion matrix counts (TP, FN, FP, TN), and performance metrics (sensitivity, specificity, PPV, and NPV). The first sheet summarizes BM-based models, and the second sheet summarizes blood-based models.

**Supplementary Table 10: Performance metrics of cfWGS vs clinical MRD calls for all models**

Performance metrics (TP, FP, SN, SP, NPV, PPV, and accuracy) for all cfWGS-based models evaluated against clinical MRD reference assays (MFC and clonoSEQ) across post-ASCT and maintenance timepoints in the training and test cohorts.

## Materials and Methods

### Patients and Study Design

A total of 51 patients were enrolled across three cohorts: (i) the Multiple Myeloma Molecular Monitoring (M4) study (IRB #17-5429, n = 41, 2018-2023) at 8 sites across Canada, (ii) the IMMAGINE and LIBERATE studies at the Princess Margaret Cancer Centre (IRB #16-5260 and #17-5357, respectively, n = 5, 2019-2023), and (iii) the Mayo Clinic Multiple Myeloma SPORE (IRB #23-5134, n = 5, 2013-2021). All patients had a baseline BM or blood cfDNA sample plus at least one BM MRD test at a later date, timed with a matching cfDNA sample. Relapse was defined per IMWG criteria.^54^

Patients were classified into a training cohort (n=44) and a test cohort (n=7). The training cohort included 41 M4 patients plus three IMMAGINE/LIBERATE patients, most with standard-of-care regimens (predominantly CyBorD and autologous stem cell transplant [ASCT]). One patient received iberdomide-bortezomib-dexamethasone. Samples were collected at diagnosis, pre-ASCT, ∼100 days post-transplant, and during maintenance (∼6-month intervals until progression or loss to follow-up). In some cases, maintenance time points were collected within a few months of a key milestone due to scheduling limitations.

The test cohort included three newly diagnosed patients from the SPORE study treated with standard frontline regimens that were not uniform to the M4 regimens (earlier-era or heterogeneous PI/IMiD combinations such as VRd/KRd), one SPORE patient who received CYCLONE (a non-standard PI + IMiD regimen including cyclophosphamide, carfilzomib, thalidomide, dexamethasone), and one relapsed/refractory SPORE patient treated with salvage therapy. The test cohort also included one newly diagnosed IMMAGINE/LIBERATE patient who received frontline T cell redirecting therapy and one relapsed/refractory IMMAGINE/LIBERATE patient. These patients formed an exploratory group to assess MRD detection in the context of advanced disease, non-standard therapies, and greater biological heterogeneity. Treatment regimens received by all patients are shown in Supplementary Table 1.

Additionally, 25 healthy controls were recruited between 2020 and 2023 at the Princess Margaret Cancer Centre through the CHARM Consortium (REB #19-6239).^55^ All participants provided written informed consent per the Declaration of Helsinki, and the relevant institutional review boards approved the protocols at each participating site.

### Clinical Data Aggregation and Processing

Clinical and laboratory data for MM patients were compiled from the M4 study cohort via the Canadian Myeloma Research Group. IMMAGINE and SPORE study data were received directly from the recruiting sites. Data were collected regarding immunoglobulin levels, serum free light chains, and the level of M-proteins. Patients were classified as high or standard cytogenetic risk based on cytogenetic results. Values were aggregated at the patient and time point levels, and duplicate entries were summarized using the mean for numeric values.

### Sample Collection and Processing

Peripheral blood (PB) was collected in Streck Cell-Free DNA BCT tubes (catalogue #218997) at all sites to stabilize cfDNA, with a target collection volume of at least 15 mL per patient. Canadian samples were shipped overnight at ambient temperature to the Princess Margaret Cancer Centre (Pugh Lab) in Toronto for plasma isolation and downstream processing. Blood samples from the Mayo Clinic were centrifuged on-site to separate the plasma fraction and then either processed locally (10 samples) or shipped frozen to the Princess Margaret Cancer Centre (8 samples) for library preparation and sequencing. Once received, samples were centrifuged at 1,900 × g for 10 minutes to separate plasma from blood cells. The plasma layer was aspirated and further centrifuged at 16,000 × g for 10 minutes at 4°C to remove cellular debris. The clarified plasma was aliquoted and stored at −80°C until cfDNA extraction. Matched normal genomic DNA was isolated from the buffy coat cellular fraction and stored at −80°C.

BM aspirates from Canadian samples (collected into EDTA tubes) were first shipped to Dalhousie University (Reiman Lab) in Saint John, NB, for CD138^+^ cell selection. CD138^+^ BM plasma cells (PCs) were enriched using magnetic bead selection (EasySep™ Human CD138 Positive Selection Kit II from Stem Cell Technologies, category #17877) to isolate tumor cells for downstream analyses. Genomic DNA from BM PCs was extracted using Qiagen DNeasy Blood and Tissue kit (250), category #69506. Yields and quality were measured by Qubit (Thermo Fisher Scientific) and/or TapeStation (Agilent). Extracted DNA was shipped to Toronto (Pugh Lab) for library preparation and sequencing.

BM and buffy coat samples from Mayo Clinic Rochester (SPORE) were processed and sequenced locally, as well as 10/18 (56%) of the cfDNA samples. Bone marrow samples were first enriched for CD138^+^ cells using anti-CD138 antibodies on a RoboSep instrument (StemCell Technologies). DNA was extracted using the Puregene kit (Qiagen).

### Library Construction and Whole-Genome Sequencing

cfDNA libraries were prepared using the KAPA HyperPrep kit (Roche) with dual indexes (Integrated DNA Technologies, IDT) from a minimum input of 20 ng of DNA. Libraries were quantified by Qubit and sequenced to 30-40X mean coverage on an Illumina NovaSeq 6000 using a 2 × 150 bp run configuration at the Ontario Institute for Cancer Research (OICR).

BM (tumor) and matched normal (buffy coat) libraries for M4 samples were prepared at the Princess Margaret Cancer Centre using 500 ng of sheared DNA (Covaris LE220), while IMMAGINE BM and germline libraries were prepared at OICR using 50 ng input. All libraries were constructed with the KAPA HyperPrep kit and sequenced on the NovaSeq 6000 at OICR. M4 tumor and germline samples were sequenced to a target depth of 30–40×. Due to unknown plasma cell content, IMMAGINE tumor samples were sequenced to 80× coverage and matched germline to 30×.

BM and buffy coat samples from the Mayo Clinic were sequenced locally on a NovaSeq6000 to 60X target coverage, with 30X target coverage for germline. In addition, 10/18 cfDNA samples from the SPORE study were sequenced locally to 30× target coverage. The remaining 8 samples were sent to OICR for sequencing.

### Alignment and Preprocessing

All WGS reads from all cohorts were aligned to the human reference genome (hg38) using the OICR *bwaMem* pipeline (v1.0.0; <u>GitHub</u>), which incorporates BWA (v0.7.12), samtools (v1.9), cutadapt (v1.8.3), and other dependencies under default parameters. Post-alignment processing included coordinate sorting (*samtools* v1.9), duplicate marking (*Picard* v2.19.2), local realignment around indels (*GATK* IndelRealigner), and base quality score recalibration (*GATK* BaseRecalibrator), as specified in the OICR *bamMergePreprocessing* workflow (v2.1.1; <u>GitHub</u>).

### Variant Calling and Annotation

Somatic single-nucleotide variants (SNVs) and small insertions/deletions (indels) were called from tumor/normal paired alignments using the OICR mutect2 pipeline (v1.0.9; <u>GitHub</u>), which uses GATK Mutect2 with samtools (v1.9). Variant annotation was performed with the OICR variantEffectPredictor workflow (v2.3.3; <u>GitHub</u>), which includes VEP (v105.0), bcftools (v1.9), bedtools (v2.27), tabix (v0.2.6), and vcf2maf (v1.6.21b). cfDNA CNAs were called with IchorCNA^53^ (hg38; autosome training; PoN; 1 Mb bins; minSegmentBins=50, txnE=0.9999, txnStrength=10000; ploidy {2,3}, normal {0.5–0.95}), yielding tumor-fraction and CN profiles. BM CNAs were called with Sequenza (v2.1.2m, γ = 400; <u>GitHub</u>); per sample we derived baseline-aware CN labels from CNt/A/B, mapped to cytoband arms. Structural variants (SVs) of immunoglobulin loci were assessed with IgCaller^56^ and custom R filtering scripts. Baseline concordance was measured by the average Jaccard index.

### Mutation-Based MRD Detection

Genomic DNA extracted from CD138^+^ BM plasma cells was subjected to WGS (≥30× target coverage) at diagnosis, thereby defining a set of baseline somatic mutations. These mutations were filtered to include only those with a VAF >10%, that passed GATK FilterMutectCalls, and did not overlap the ENCODE Blacklist v2^57^ or the UCSC hg38 RepeatMasker annotation (RepeatMasker v4.1.6, hg38.fa.out.gz, downloaded from the UCSC Genome Browser).^58^ As a conservative measure to avoid germline polymorphisms, we also excluded all variants annotated with an rsID, acknowledging that this may remove a small number of true somatic events.

To generate mutation lists from baseline blood cfDNA for tumor-naïve MRD detection, we applied the same filtering approach as before, with one modification in that mutations were retained if supported by at least two reads, rather than using a strict VAF threshold (>10%). This strategy was designed to reduce likely sequencing artifacts while retaining low-VAF mutations present in baseline cfDNA.

We defined a baseline mutation list as high quality if the sample showed evidence of disease at diagnosis, meaning it contained an IGH driver, a highest-VAF SNV of at least 10% (or at least 5% for cfDNA), a tumor fraction of at least 10% in BM or at least 5% in cfDNA, or a cfDNA tumor fraction of 3–5% with a myeloma-specific copy-number abnormality (del1p, amp1q, del13q, or del17p). Baseline cfDNA samples that did not meet these criteria were still classified as high quality if their filtered mutation count exceeded the first-quartile threshold of the mutation-count distribution among evidence-positive samples. These high-quality mutation lists were subsequently tracked in longitudinal cfDNA samples across treatment and maintenance time points in addition to fragmentomic features. Mutation tracking in cfDNA was performed using the MRDetect algorithm (as in Zviran et al.^30^), available by an academic license from the authors. For each cfDNA sample, we calculated several metrics as defined below:

*Sites checked*

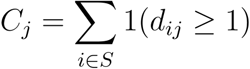

This counts how many baseline tumor mutations had at least one read covering them in sample *j*. Here, *i* indexes each mutation in the baseline set *S*, *j* indexes the cfDNA sample, and *d_ij_* is the total number of reads covering locus *i* in sample *j*.

*Sites detected*

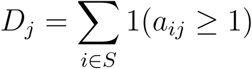

This counts how many of the checked sites show evidence of the mutant allele. Here, *a_ij_* is the number of reads supporting the alternate allele at locus *i* in sample *j*.

*Proportion of sites detected*

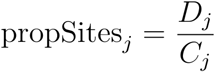

This represents the fraction of checked sites that are observed with mutant evidence.

*Cumulative VAF (cVAF)*

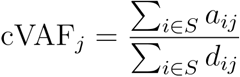

This measures the pooled proportion of mutant reads out of all reads across the baseline mutation sites.

#### Z-scores relative to controls

To distinguish true tumor signal from background noise, the metrics *x_j_* ε {propSites*_j_*, cVAF*_j_*}. are standardized relative to 25 healthy controls. The standardized score is defined as

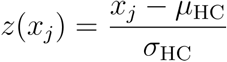

Where μ_HC_ and σ_HC_ are the mean and standard deviation of the metric across healthy control samples. A large positive z-score indicates significantly higher signal than expected from background error rates.

### Clinical MRD Comparators

cfWGS MRD was evaluated against BM MFC MRD and/or BM targeted sequencing of immunoglobulin loci (clonoSeq).^59^ MFC was conducted within 48 hours of bone marrow sample collection at either Cytoquest Corporation (Toronto, ON) or the Flow Cytometry Division at Toronto General Hospital. Samples were analyzed on a FACSCanto II flow cytometer (BD Biosciences, San Jose, CA) using an eight-color, two-tube lyse/wash assay that included intracellular staining for cytoplasmic kappa and lambda chains, alongside surface markers CD38, CD27, CD138, CD56, CD45, CD19, CD117, and CD81. Data acquisition and analysis followed EuroFlow guidelines, and MRD was reported as negative if fewer than 20 aberrant plasma cells were detected within ≥2,000,000 acquired events, achieving a target sensitivity of at least 10^-5^.^60^

Targeted immunoglobulin sequencing for MRD was done with the clonoSEQ assay (Adaptive Biotechnologies)^61^, an FDA-cleared in vitro diagnostic test that detects and quantifies clonotypic immunoglobulin (IgH, IgK, IgL) rearrangements in BM genomic DNA. At diagnosis, dominant clonotypes meeting predefined criteria were identified and subsequently tracked in follow-up samples. MRD was reported quantitatively as a per-million count of malignant cells, with sensitivity thresholds of 10^-5^ to 10^-6^, depending on DNA input quantity.

### Fragmentomics Analysis

Fragmentomic features were derived with the Pugh Lab *pipeline-suite* fragmentomics workflow (v0.9.11) using hg38 (<u>Github</u>). To control technical variation from library size and stabilize variance in length-based metrics, BAMs were downsampled to 50 million uniquely mapped, concordantly paired-end reads per sample before feature extraction. PCR duplicates were removed with Picard (v2.10.9). Insert size was then evaluated using Picard CollectInsertSizeMetrics and the proportion of short fragments was calculated as the ratio of 10-150bp fragments divided by the total cfDNA fragments (10-600bp). A weighted fragment length score derived from global insert size distributions relative to healthy controls was then calculated as described by Vessies et al.^61^ using default tool parameters.

To profile regional chromatin accessibility, we ran Griffin (v0.1.0) with GC-bias modeling/correction and mappability filtering (Umap k-50), using as sites the 1,556 de novo active H3K27ac-marked regulatory regions gained in MM and absent in normal B-cell differentiation, as defined in Ordoñez *et al.*^62^ Griffin site profiles were summarized over a ∼±1 kb window centered on each regulatory element (positions −990 to +975, 15 bp bins). Mean coverage was defined as the arithmetic mean of GC-corrected per-bin coverage across this window, with midpoint coverage defined as the average at −30/−15/0/+15/+30 bp and amplitude as the periodogram maximum. This analysis aimed to assess whether incorporating fragmentomic features could improve detection sensitivity and accuracy beyond mutation-based approaches.

### Feature Integration and Classifier Optimization for MRD Detection

To develop a classifier for cfDNA-based MRD detection, we defined a library of candidate feature combinations derived from cfDNA and BM sequencing data. The candidate features included both single-feature models and multi-feature combinations. The single-feature models included the “sites” model, defined as the z-score of the proportion of baseline-identified mutation loci detected in cfDNA relative to healthy controls, and the “cVAF” model, defined as the z-score of the cumulative variant allele fraction (cVAF) across those loci relative to healthy controls. The multi-feature models integrated additional fragmentomic and genomic metrics, including a fragment size score,^63^ mean sequencing coverage at MM-specific enhancer regions previously identified by ATAC-Seq,^62^ the proportion of short cfDNA fragments, and tumor fraction estimated by ichorCNA.

We trained and evaluated models separately for BM-derived and blood-derived cfDNA predictors, as well as combined models incorporating both signals. All model training was performed using data from the training induction–transplant cohort, which served as the training set. The test cohort was held out entirely and used exclusively for independent testing and validation. Model training used elastic net-penalized logistic regression implemented using the glmnet package (v4.1-8), with hyperparameter tuning across a grid of alpha (elastic net mixing parameter: 0, 0.25, 0.5, 0.75, 1) and lambda (regularization strength, spanning 50 log-scaled values from 0.001 to 10). Single-feature models were fit using standard logistic regression (glm) without penalization. All features were centred and scaled using the caret package (v7.0.1).

Model optimization and internal validation were performed using a nested cross-validation (CV) strategy. A five-fold stratified outer CV loop was used to estimate model performance while minimizing overfitting. For each outer fold, we performed a 5×5 repeated inner CV to determine optimal hyperparameters. Model performance metrics, including area under the receiver operating characteristic curve (AUC), sensitivity, specificity, balanced accuracy, positive predictive value (PPV), negative predictive value (NPV), F₁ score, Brier score, and partial AUC (pAUC90) focused on high-specificity regions (≥90%), were computed from the out-of-fold predictions. These metrics were averaged across folds to generate robust generalization estimates. Following the nested CV, the best-performing feature combinations were selected and refitted on the entire training set to establish final model coefficients and fixed decision thresholds based on the Youden index (maximizing balanced accuracy). The resulting classifiers were then applied to the rest of the cohort and dilution series to assess generalizability and limits of detection.

### Dilution Series Experiment

To evaluate the theoretical limits of detection for cfDNA-based MRD approaches, we performed a dilution series experiment that mixed cfDNA libraries from two time points from a single MM patient. Specifically, we combined a baseline cfDNA library (aggregate VAF = 0.688%) with a cfDNA library from blood collected at an MRD-negative, post-induction and transplant time point (VAF = 0.0087%, determined from the cumulative VAF of all BM somatic mutations detected in cfDNA by cfWGS). To target progressively decreasing mutant fractions, we combined these libraries at ratios of 100%, 61.6%, 30.8%, 15.4%, 7.7%, 3.9%, 2.0%, 0.3%, and 0% baseline library (with the remainder post-induction and transplant library). These correspond to cVAF levels of 0.688% (no post-induction and transplant library), 0.427%, 0.218%, 0.113%, 0.061%, 0.035%, 0.022%, 0.011%, and 0.0087% (no diagnostic cfDNA library). To establish the ctDNA content of the diagnostic sample, we first identified somatic mutations from WGS of BM-derived CD138^+^ cells. We then calculated the aggregate VAF in the baseline cfDNA by summing the variant allele fractions of all tumor-identified mutations from the bone marrow sample that had a VAF greater than 10%. Each combination of library dilution was sequenced to 40X WGS coverage using an Illumina NovaSeq 6000 using a 2 × 150 bp run configuration at OICR. We then extracted 7 features (detection rate as reads detected / reads checked, z-scores of the detection rate and proportion of sites detected, fragmentomic metrics, classifier probabilities) and assessed monotonic association using Spearman correlation between each feature and the estimated limit of detection to empirically determine the theoretical sensitivity and lower limit of detection achievable by our method.

### Statistical Analysis

Comparisons among MRD assays were performed using Pearson or Spearman correlation (for continuous variables) and Cohen’s kappa (for categorical positivity/negativity). Contingency tables and scatter plots further assessed concordance across methods as appropriate. Clinical correlation, including association with PFS or OS, was tested via Kaplan–Meier curves and log-rank tests. PFS was defined as the time from each landmark blood draw (100 days post-ASCT or 1 year on maintenance) to the first of either IMWG-defined progression or death from any cause. Patients alive and progression-free at last contact were censored at their last follow-up date. Cox proportional hazards regression models adjusted for known covariates were fitted to estimate hazard ratios. All statistical analyses were performed using R (v4.3.3), and p<0.05 (two-sided) was considered significant.

### Data and Code Availability

Primary sequence data that support the findings of this study will be available under controlled access from the European Genome-Phenome Archive (EGA, upload in progress). All secondary data derived from the primary alignments, custom scripts to reproduce the MRD analyses and relevant pipelines are available at https://github.com/pughlab/cfWGS-MM-MRD.

## Supplementary Notes

### Baseline Genomic Characteristics of Cohort

Concordance between matched BM and cfDNA at diagnosis, measured by the average Jaccard index across patients, was high for translocations (training mean 0.98, test mean 0.86) but lower for CNAs (0.45 vs 0.64) and mutations (0.59 vs 0.33) (Supplementary Table 2). As circulating tumour DNA burden can affect variant recovery, we next examined tumour fraction in plasma. The median tumor fraction estimate was 5.1% (range 0-33.7%) in the training and 5.3% (0-18.2%) in the test cohort, with ≥5% ctDNA in 51% and 60% of samples, respectively (Supplementary Figure 2A). Plasma tumour fraction had minimal impact on overall concordance for structural variants and CNAs (Supplementary Figure 2B). In contrast, sensitivity for detecting BM-derived mutations in cfDNA was higher in samples with high tumour fraction (defined as ≥5% ctDNA fraction; 21% vs 0%), with a similar pattern for CNAs (77% vs 33%). Sensitivity for translocations was 50% in high-TF samples, although numbers were small (Supplementary Figure 2B; Supplementary Table 2A).

To further evaluate the performance of WGS-based structural variant (SV) detection in clinical samples, we next compared WGS calls to orthogonal results from FISH. This analysis aimed to assess the sensitivity and reliability of SV detection in both BM-derived DNA and cfDNA. In the training cohort, BM WGS agreed with FISH in 83% of translocation calls and 79% of CNAs; corresponding cfDNA concordance was 82% and 88% (Supplementary Figure 2C). However, most translocation concordance was driven by cases in which both assays reported no detectable event, and sensitivity to detect translocations was only 25% in BM and 10% by baseline cfDNA using WGS (Supplementary Figure 2C, Supplementary Table 2). CNA detection showed moderate sensitivity to FISH, although event counts were small for individual alterations. FISH identified amp(1q), del(17p), and del(1p). BM WGS confirmed amp(1q) in 1 of 2 FISH-positive cases, while the second case, with ploidy of 4, likely reflected a ploidy-related gain rather than a true amplification. BM WGS also detected a focal 1q gain slightly distal to the FISH probe position (Supplementary Figure 2C-D, Supplementary Table 2). Del(17p) was confirmed in 2 of 4 cases and del(1p) in 1 of 1. In plasma cfWGS, sensitivity relative to FISH was 80% for amp(1q) (4/5), with the remaining case representing the same focal 1q gain identified in BM, detected distal to the FISH probe in plasma as well, but below arm-level thresholds in WGS. Sensitivity for del(17p) was 43% (3/7), with detection in 3/5 high–tumor-fraction samples and none of the low–tumor-fraction samples, while del(1p) was concordantly detected (1/1). These results highlight the influence of probe localization and ploidy on 1q concordance and underscore the dependence of del(17p) sensitivity on tumor fraction (Supplementary Figure 2C, Supplementary Table 2).

To enable personalized MRD tracking, we next profiled baseline somatic mutations in BM and cfDNA and assessed their relationship to clinical and fragmentomic features. In total, we detected an average of 2,400 mutations per baseline BM in the training cohort (range: 106–8,721, SD = 1,701) and 2,287 per baseline cfDNA (range: 652–7,323, SD = 1,217) (Supplementary Figure 2E). In the test cohort, a significantly higher number of mutations were identified in cfDNA (p < 0.01, Wilcoxon rank-sum), with an average of 4,311 mutations (range: 2,554–6,273, SD = 1,492) compared to training cohort cases. While a similar trend was observed in BM samples (average: 2,998 mutations, range: 1,102–4,432, SD = 1,095), the difference did not reach statistical significance (p = 0.13) (Supplementary Figure 2E). The higher mutation counts in the test cohort likely reflect the inclusion of relapsed/refractory and more cytogenetically defined high-risk patients, which are often associated with greater genomic complexity and higher ctDNA burden.^1–3^ However, the small cohort size may also exaggerate this difference. While mutation counts in our study were lower than those reported in some previous WGS analyses, such as CoMMpass^4^ and Oben et al.^5^, this likely reflects differences in filtering approaches. Specifically, our use of conservative thresholds (e.g., VAF ≥10% and filtering of regions annotated by RepeatMasker^6^) was intended to prioritize high-confidence, clonal mutations suitable for MRD tracking.

We observed a moderate positive correlation between baseline BM and cfDNA mutation counts (Spearman ρ = 0.40, p = 0.025; Supplementary Figure 2F). In cfDNA, mutation count also correlated with the CNA-derived tumor fraction (from ichorCNA) (ρ = 0.52, p < 0.01), the proportion of short-fragment score (ρ = 0.31, p = 0.03), and was inversely correlated with serum albumin (ρ = –0.37, p = 0.02; Supplementary Figure 2F). After Benjamini–Hochberg correction, only the cfDNA mutation count–CNA-derived tumor fraction association remained significant (adjusted p < 0.01; Supplementary Figure 2F, Supplementary Table 3). Collectively, these results highlight the feasibility of baseline genomic profiling from cfDNA and BM samples, substantial yet variable concordance between sample types, and strong relationships between cfDNA mutation burden and both tumor fraction and fragmentomic features.

### Longitudinal cfDNA-based MRD Analysis

Longitudinal analyses were performed in the training cohort (n = 44) to characterize cfDNA dynamics during therapy. Across the series (144 cfDNA draws with ≥1 feature from 44 patients; median follow-up 25 months, range 4–49), cfDNA features followed treatment response (Figure 2A). For subset analyses, BM-derived features were calculated from 95 draws in 27 patients with disease-associated alterations detected in BM at diagnosis (median follow-up 25 months, range 4–49); blood-derived features from 102 draws in 30 patients with disease-associated alterations detected in blood at diagnosis (median 25 months, range 7–47); and fragmentomic features from all 144 draws. We evaluated three metrics from baseline mutation lists: (i) z-score of the proportion of mutant sites detected versus healthy, (ii) z-score of cumulative VAF (cVAF), and (iii) raw cVAF (**Methods)**. With BM-derived mutation lists, the site-detection z-score declined from a median of 130.07 to 0.44 and the cVAF z-score from a median of 477.1 to 2.35 from diagnosis to first on-treatment time point (both fold change <0.01×; p <0.001); the cVAF declined by 3.43 percentage points (median 3.43% to 0.05%; fold change = 0.02×; p = <0.001; Figure 2B, Supplementary Figure 3A). These metrics correlated with clinical biomarkers, including M-protein levels (ρ = 0.74), the percentage of aberrant plasma cells by MFC (ρ = 0.82), and the frequency of the dominant immunoglobulin rearrangement by clonoSEQ (ρ = 0.83), all with BH-adjusted p-values < 0.05 (Supplementary Figure 4, Supplementary Table 3). BM-derived metrics dropped sharply after therapy and tracked clinical biomarkers, supporting their use as dynamic measures of disease burden.

Using blood-derived baseline lists, trends were concordant despite modest BM–blood mutation overlap (mean 11%, range 0.04–49.4%, interquartile range [IQR] = 0.6-17.1%) (Supplementary Figure 2G). The site-detection z-score declined from a median of 97.54 to 10.82 (fold change = 0.11×; p = <0.001), the cVAF z-score from 59.18 to 8.41 (fold = 0.14×; p = <0.001), and the raw cVAF from 14.37% to 3.16% (fold = 0.21×; p = <0.001) following treatment initiation (Figure 2C, Supplementary Figure 3B). Blood-derived metrics were strongly correlated with their BM counterparts. For instance, the cVAF z-scores in blood and BM were significantly correlated (ρ = 0.76, p < 0.001), as were the site-detection z-scores (ρ = 0.85, p < 0.001; Supplementary Figure 4). Blood-derived metrics were also significantly correlated with clinical biomarkers. For example, the cVAF z-score showed significant correlations with M-protein levels (ρ = 0.64), the κ:λ light chain ratio (ρ = 0.34), the percentage of aberrant plasma cells by MFC (ρ = 0.68), and the frequency of the dominant immunoglobulin rearrangement by clonoSEQ (ρ = 0.71), all with BH-adjusted p-values < 0.05 (Supplementary Figure 4, Supplementary Table 3). When baseline mutation lists were derived from peripheral-blood cfDNA, longitudinal metrics closely recapitulated those from BM and clinical markers, supporting the feasibility of blood cfDNA-only designs when BM is unavailable.

Fragmentomic signals moved in tandem from baseline to first treatment. The fragment-size score contracted by 25.65%, indicating an increasing proportion of long fragments (p = <0.001), the mean coverage over MM enhancers increased marginally from 0.98× to 0.99×, indicating less active transcription at these sites (p = <0.01), and the proportion of short fragments decreased from 17.53% to 14.82% (fold change = 0.85×; p = <0.01) (Figure 2D). Meanwhile, ichorCNA tumour fraction dropped from a median of 5.1% to 3.4% (fold change = 0.68×; p = <0.001; Figure 2D, Supplementary Figure 3C), which is below the quantitative range of this tool, and few high-quality CNVs calls were discernible. Multiple fragmentomic features were significantly correlated with the frequency of the dominant immunoglobulin rearrangement by clonoSEQ, including the fragment size score (ρ = 0.42), proportion of short fragments (ρ = 0.35), coverage at MM transcription sites (ρ = -0.51), and tumor fraction (ρ = 0.43), all with BH-adjusted p-values < 0.05. These correlations were comparable in strength to those observed with the percentage of aberrant plasma cells in bone marrow by MFC (Supplementary Figure 4, Supplementary Table 3). Fragmentomic features track response and correlate with immunoglobulin-based burden, supporting their use as complementary signals alongside mutation metrics.

Together, these results demonstrate that both mutation-based and fragmentomic cfDNA features robustly capture therapy-aligned disease dynamics in newly diagnosed patients. Motivated by these insights, we next evaluated the performance of single- and multi-feature classifiers for cfDNA-based MRD detection.

## Notes

### Author Declarations

The study was approved by the Research Ethics Boards of all participating institutions, including the Princess Margaret Cancer Centre (University Health Network; IRB #16-5260, #17-5357, #19-6239), the Canadian M4 study (multi-centre IRB #17-5429), and the Mayo Clinic (IRB #23-5134). All participants provided written informed consent in accordance with the Declaration of Helsinki.

## References

1. Rajkumar, S. V. Multiple myeloma: 2024 update on diagnosis, risk-stratification, and management. Am. J. Hematol. 99, 1802–1824 (2024).

2. Pan, T., Zhang, J., Wang, X. & Song, Y. Global burden and trends of hematologic malignancies based on Global Cancer Observatory 2022 and Global Burden of Disease 2021. Exp. Hematol. Oncol. 14, 98 (2025).

3. Voorhees, P. M. et al. Addition of daratumumab to lenalidomide, bortezomib, and dexamethasone for transplantation-eligible patients with newly diagnosed multiple myeloma (GRIFFIN): final analysis of an open-label, randomised, phase 2 trial. Lancet Haematol. 10, e825–e837 (2023).

4. Palumbo, A. et al. Daratumumab, Bortezomib, and Dexamethasone for Multiple Myeloma. N. Engl. J. Med. 375, 754–766 (2016).

5. Kumar, S. K. et al. Continued improvement in survival in multiple myeloma: changes in early mortality and outcomes in older patients. Leukemia 28, 1122–1128 (2014).

6. Moreau, P. et al. Maintenance with daratumumab or observation following treatment with bortezomib, thalidomide, and dexamethasone with or without daratumumab and autologous stem-cell transplant in patients with newly diagnosed multiple myeloma (CASSIOPEIA): an open-label, randomised, phase 3 trial. Lancet Oncol. 22, 1378–1390 (2021).

7. Munshi, N. C. et al. A large meta-analysis establishes the role of MRD negativity in long-term survival outcomes in patients with multiple myeloma. Blood Adv. 4, 5988–5999 (2020).

8. Cavo, M. et al. Prognostic value of minimal residual disease negativity in myeloma: combined analysis of POLLUX, CASTOR, ALCYONE, and MAIA. Blood 139, 835–844 (2022).

9. Kostopoulos, I. V., Ntanasis-Stathopoulos, I., Gavriatopoulou, M., Tsitsilonis, O. E. & Terpos, E. Minimal Residual Disease in Multiple Myeloma: Current Landscape and Future Applications With Immunotherapeutic Approaches. Front. Oncol. 10, 860 (2020).

10. Landgren, O. & Devlin, S. M. Minimal Residual Disease as an Early Endpoint for Accelerated Drug Approval in Myeloma: A Roadmap. Blood Cancer Discov. 6, 13 (2024).

11. Medina-Herrera, A., Sarasquete, M. E., Jiménez, C., Puig, N. & García-Sanz, R. Minimal Residual Disease in Multiple Myeloma: Past, Present, and Future. Cancers 15, 3687 (2023).

12. Avet-Loiseau, H. et al. Minimal Residual Disease Status as a Surrogate Endpoint for Progression-free Survival in Newly Diagnosed Multiple Myeloma Studies: A Meta-analysis. Clin. Lymphoma Myeloma Leuk. 20, e30–e37 (2020).

13. Perrot, A. et al. Minimal residual disease negativity using deep sequencing is a major prognostic factor in multiple myeloma. Blood 132, 2456–2464 (2018).

14. Rasche, L. et al. Spatial genomic heterogeneity in multiple myeloma revealed by multi-region sequencing. Nat. Commun. 8, 268 (2017).

15. Derlin, T. et al. 18F-FDG PET/CT for detection and localization of residual or recurrent disease in patients with multiple myeloma after stem cell transplantation. Eur. J. Nucl. Med. Mol. Imaging 39, 493–500 (2012).

16. Jamet, B. et al. Functional Imaging for Therapeutic Assessment and Minimal Residual Disease Detection in Multiple Myeloma. Int. J. Mol. Sci. 21, 5406 (2020).

17. Puig, N. et al. Reference Values to Assess Hemodilution and Warn of Potential False-Negative Minimal Residual Disease Results in Myeloma. Cancers 13, 4924 (2021).

18. Noori, S. et al. Dynamic monitoring of myeloma minimal residual disease with targeted mass spectrometry. Blood Cancer J. 13, 30 (2023).

19. Langerhorst, P. et al. Multiple Myeloma Minimal Residual Disease Detection: Targeted Mass Spectrometry in Blood vs Next-Generation Sequencing in Bone Marrow. Clin. Chem. 67, 1689–1698 (2021).

20. Ye, X., Li, W., Zhang, L. & Yu, J. Clinical Significance of Circulating Cell-Free DNA Detection in Multiple Myeloma: A Meta-Analysis. Front. Oncol. 12, (2022).

21. Wan, J. C. M. et al. Liquid biopsies come of age: towards implementation of circulating tumour DNA. Nat. Rev. Cancer 17, 223–238 (2017).

22. Gerber, B. et al. Circulating tumor DNA as a liquid biopsy in plasma cell dyscrasias. Haematologica 103, e245–e248 (2018).

23. Chow, S. et al. Myeloma immunoglobulin rearrangement and translocation detection through targeted capture sequencing. Life Sci. Alliance 6, (2023).

24. Kis, O. et al. Circulating tumour DNA sequence analysis as an alternative to multiple myeloma bone marrow aspirates. Nat. Commun. 8, 1–11 (2017).

25. Guo, G. et al. Genomic discovery and clonal tracking in multiple myeloma by cell free DNA sequencing. Leukemia 32, 1838 (2018).

26. Waldschmidt, J. M. et al. Cell-free DNA for the detection of emerging treatment failure in relapsed/refractory multiple myeloma. Leukemia 36, 1078–1087 (2022).

27. Hosoya, H. et al. Deciphering response dynamics and treatment resistance from circulating tumor DNA after CAR T-cells in multiple myeloma. Nat. Commun. 16, 1824 (2025).

28. Walker, B. A. et al. Intraclonal heterogeneity and distinct molecular mechanisms characterize the development of t(4;14) and t(11;14) myeloma. Blood 120, 1077–1086 (2012).

29. Miller, A. et al. High somatic mutation and neoantigen burden are correlated with decreased progression-free survival in multiple myeloma. Blood Cancer J. 7, e612–e612 (2017).

30. Zviran, A. et al. Genome-wide cell-free DNA mutational integration enables ultra-sensitive cancer monitoring. Nat. Med. 26, 1114–1124 (2020).

31. Cristiano, S. et al. Genome-wide cell-free DNA fragmentation in patients with cancer. Nature 570, 385–389 (2019).

32. Parsons, H. A. et al. Sensitive Detection of Minimal Residual Disease in Patients Treated for Early-Stage Breast Cancer. Clin. Cancer Res. 26, 2556–2564 (2020).

33. Wang, S. et al. Enhanced Detection of Landmark Minimal Residual Disease in Lung Cancer Using Cell-free DNA Fragmentomics. Cancer Res. Commun. 3, 933–942 (2023).

34. Haider, Z. et al. Whole-genome informed circulating tumor DNA analysis by multiplex digital PCR for disease monitoring in B-cell lymphomas: a proof-of-concept study. Front. Oncol. 13, (2023).

35. Ding, S. C. & Lo, Y. M. D. Cell-Free DNA Fragmentomics in Liquid Biopsy. Diagn. Basel Switz. 12, 978 (2022).

36. van’t Erve, I., et al. Cancer treatment monitoring using cell-free DNA fragmentomes. Nat. Commun. 15, 8801 (2024).

37. Widman, A. J. et al. Ultrasensitive plasma-based monitoring of tumor burden using machine-learning-guided signal enrichment. Nat. Med. 30, 1655–1666 (2024).

38. Usmani, S. Z. et al. Daratumumab plus bortezomib, lenalidomide and dexamethasone for transplant-ineligible or transplant-deferred newly diagnosed multiple myeloma: the randomized phase 3 CEPHEUS trial. Nat. Med. 31, 1195–1202 (2025).

39. Sonneveld, P. et al. Daratumumab, Bortezomib, Lenalidomide, and Dexamethasone for Multiple Myeloma. N. Engl. J. Med. 390, 301–313 (2024).

40. Moreau, P. et al. Bortezomib, thalidomide, and dexamethasone with or without daratumumab before and after autologous stem-cell transplantation for newly diagnosed multiple myeloma (CASSIOPEIA): a randomised, open-label, phase 3 study. Lancet Lond. Engl. 394, 29–38 (2019).

41. Costa, L. J. et al. Daratumumab, Carfilzomib, Lenalidomide, and Dexamethasone With Minimal Residual Disease Response-Adapted Therapy in Newly Diagnosed Multiple Myeloma. J. Clin. Oncol. Off. J. Am. Soc. Clin. Oncol. 40, 2901–2912 (2022).

42. Puig, N. et al. Measurable residual disease by mass spectrometry and next-generation flow to assess treatment response in myeloma. Blood 144, 2432–2438 (2024).

43. Lasa, M. et al. Minimally Invasive Assessment of Peripheral Residual Disease During Maintenance or Observation in Transplant-Eligible Patients With Multiple Myeloma. J. Clin. Oncol. 43, 125–132 (2025).

44. Firestone, R. S. et al. Antigen escape as a shared mechanism of resistance to BCMA-directed therapies in multiple myeloma. Blood 144, 402–407 (2024).

45. Lee, H. et al. Mechanisms of antigen escape from BCMA- or GPRC5D-targeted immunotherapies in multiple myeloma. Nat. Med. 29, 2295–2306 (2023).

46. Jiang, P. et al. Plasma DNA End-Motif Profiling as a Fragmentomic Marker in Cancer, Pregnancy, and Transplantation. Cancer Discov. 10, 664–673 (2020).

47. Mathios, D. et al. Detection and characterization of lung cancer using cell-free DNA fragmentomes. Nat. Commun. 12, 5060 (2021).

48. Vanderstichele, A. et al. Nucleosome footprinting in plasma cell-free DNA for the pre-surgical diagnosis of ovarian cancer. Npj Genomic Med. 7, 1–9 (2022).

49. Li, X. et al. Ultra-sensitive molecular residual disease detection through whole genome sequencing with single-read error correction. EMBO Mol. Med. 16, 2188–2209 (2024).

50. Martello, M. et al. High level of circulating cell-free tumor DNA at diagnosis correlates with disease spreading and defines multiple myeloma patients with poor prognosis. Blood Cancer J. 14, 208 (2024).

51. Roschewski, M. et al. Circulating tumour DNA and CT monitoring in patients with untreated diffuse large B-cell lymphoma: a correlative biomarker study. Lancet Oncol. 16, 541–549 (2015).

52. Williams, M. J. et al. Tracking clonal evolution during treatment in ovarian cancer using cell-free DNA. Nature 1–9 (2025) doi:10.1038/s41586-025-09580-0.

53. Adalsteinsson, V. A. et al. Scalable whole-exome sequencing of cell-free DNA reveals high concordance with metastatic tumors. Nat. Commun. 2017 81 8, 1–13 (2017).

54. Kumar, S. et al. International Myeloma Working Group consensus criteria for response and minimal residual disease assessment in multiple myeloma. Lancet Oncol. 17, e328–e346 (2016).

55. Farncombe, K. M. et al. Current and new frontiers in hereditary cancer surveillance: Opportunities for liquid biopsy. Am. J. Hum. Genet. 110, 1616–1627 (2023).

56. Medina, A. et al. Molecular profiling of immunoglobulin heavy-chain gene rearrangements unveils new potential prognostic markers for multiple myeloma patients. Blood Cancer J. 2020 102 10, 1–12 (2020).

57. Amemiya, H. M., Kundaje, A. & Boyle, A. P. The ENCODE Blacklist: Identification of Problematic Regions of the Genome. Sci. Rep. 9, 9354 (2019).

58. Dfam-consortium/RepeatMasker. Dfam Consortium (2025).

59. Faham, M. et al. Deep-sequencing approach for minimal residual disease detection in acute lymphoblastic leukemia. Blood 120, 5173–5180 (2012).

60. Kalina, T. et al. EuroFlow standardization of flow cytometer instrument settings and immunophenotyping protocols. Leukemia 26, 1986–2010 (2012).

61. Ching, T. et al. Analytical evaluation of the clonoSEQ Assay for establishing measurable (minimal) residual disease in acute lymphoblastic leukemia, chronic lymphocytic leukemia, and multiple myeloma. BMC Cancer 20, 612 (2020).

62. Ordoñez, R. et al. Chromatin activation as a unifying principle underlying pathogenic mechanisms in multiple myeloma. Genome Res. 30, 1217–1227 (2020).

63. Vessies, D. C. L. et al. Combining variant detection and fragment length analysis improves detection of minimal residual disease in postsurgery circulating tumour DNA of stage II– IIIA NSCLC patients. Mol. Oncol. 16, 2719–2732 (2022).

64. Hoang, P. H. et al. Whole-genome sequencing of multiple myeloma reveals oncogenic pathways are targeted somatically through multiple mechanisms. Leukemia 32, 2459 (2018).

65. Oben, B. et al. Whole-genome sequencing reveals progressive versus stable myeloma precursor conditions as two distinct entities. Nat. Commun. 12, 1861 (2021).

## References for Supplementary Notes

1. Rasche, L. et al. Spatial genomic heterogeneity in multiple myeloma revealed by multi-region sequencing. Nat. Commun. 8, 268 (2017).

2. Martello, M. et al. High level of circulating cell-free tumor DNA at diagnosis correlates with disease spreading and defines multiple myeloma patients with poor prognosis. Blood Cancer J. 14, 208 (2024).

3. Ye, X., Li, W., Zhang, L. & Yu, J. Clinical Significance of Circulating Cell-Free DNA Detection in Multiple Myeloma: A Meta-Analysis. Front. Oncol. 12, (2022).

4. Hoang, P. H. et al. Whole-genome sequencing of multiple myeloma reveals oncogenic pathways are targeted somatically through multiple mechanisms. Leukemia 32, 2459 (2018).

5. Oben, B. et al. Whole-genome sequencing reveals progressive versus stable myeloma precursor conditions as two distinct entities. Nat. Commun. 12, 1861 (2021).

6. Dfam-consortium/RepeatMasker. Dfam Consortium (2025).

